# Computational Phenomapping of Randomized Clinical Trials to Enable Assessment of their Real-world Representativeness and Personalized Inference

**DOI:** 10.1101/2024.05.15.24306285

**Authors:** Phyllis M. Thangaraj, Evangelos K. Oikonomou, Lovedeep S. Dhingra, Arya Aminorroaya, Rahul Jayaram, Marc A. Suchard, Rohan Khera

**Affiliations:** Section of Cardiovascular Medicine, Department of Internal Medicine, Yale School of Medicine, New Haven, CT, USA; Department of Biostatistics, Fielding School of Public Health, University of California, 650 Charles E. Young Drive S, Los Angeles, CA 90095, USA; Departments of Computational Medicine and Human Genetics, David Geffen School of Medicine at UCLA, University of California, 695 Charles E. Young Drive S, Los Angeles, CA 90095, USA; Section of Health Informatics, Department of Biostatistics, Yale School of Public Health, New Haven, CT; Section of Biomedical Informatics and Data Science, Yale School of Medicine, New Haven, CT; Center for Outcomes Research and Evaluation, Yale-New Haven Hospital, New Haven, CT, USA

## Abstract

**BACKGROUND:** Randomized clinical trials (RCTs) define evidence-based medicine, but quantifying their generalizability to real-world patients remains challenging. We propose a multidimensional approach to compare individuals in RCT and electronic health record (EHR) cohorts by quantifying their representativeness and estimating real-world effects based on individualized treatment effects (ITE) observed in RCTs.

**METHODS:** We identified 65 pre-randomization characteristics of an RCT of heart failure with preserved ejection fraction (HFpEF), the Treatment of Preserved Cardiac Function Heart Failure with an Aldosterone Antagonist Trial (TOPCAT), and extracted those features from patients with HFpEF from the EHR within the Yale New Haven Health System. We then assessed the real-world generalizability of TOPCAT by developing a multidimensional machine learning-based phenotypic distance metric between TOPCAT stratified by region including the United States (US) and Eastern Europe (EE) and EHR cohorts. Finally, from the ITE identified in TOPCAT participants, we assessed spironolactone benefit within the EHR cohorts.

**RESULTS:** There were 3,445 patients in TOPCAT and 8,121 patients with HFpEF across 4 hospitals. Across covariates, the EHR patient populations were more similar to each other than the TOPCAT-US participants (median SMD 0.065, IQR 0.011-0.144 vs median SMD 0.186, IQR 0.040-0.479). At the multi-variate level using the phenotypic distance metric, our multidimensional similarity score found a higher generalizability of the TOPCAT-US participants to the EHR cohorts than the TOPCAT-EE participants. By phenotypic distance, a 47% of TOPCAT-US participants were closer to each other than any individual EHR patient. Using a TOPCAT-US-derived model of ITE from spironolactone, all patients were predicted to derive benefit from spironolactone treatment in the EHR cohort, while a TOPCAT-EE-derived model predicted 13% of patients to derive benefit.

**CONCLUSIONS:** This novel multidimensional approach evaluates the real-world representativeness of RCT participants against corresponding patients in the EHR, enabling the evaluation of an RCT’s implication for real-world patients.

## INTRODUCTION

Randomized clinical trials (RCTs) are the standard for defining optimal care practices, but quantifying their generalizability to real-world patients remains challenging.^1–4^ Underrepresentation and under-enrollment of key patient demographic and clinical subpopulations contribute to this gap, leading to decreased external validity of RCT treatment effect outcomes in these populations. ^5–12^ The generalizability of RCTs across real-world populations relies on their external validity^3^, however, prior studies have been unable to capture the complete profile of patients, relying instead on comparing populations across a few covariates or one covariate at a time.^12–15^ For example, hypothetically, if an RCT had an equal gender distribution, but all men had renal disease, and all women had diabetes, a real-world cohort with similar gender composition but with renal disease and diabetes present split equally between genders would be indistinguishable on univariate comparisons of gender, diabetes, or renal dysfunction. This example, while an extreme case, does not even account for the complex relationships across all covariates. Therefore, a multi-dimensional phenotypic representation of cohorts is needed to adequately evaluate representativeness between RCT cohorts and real-world populations.

To address this, we leveraged participant-level data from a large, phase 3 RCT of heart failure with preserved ejection fraction (HFpEF).^16,17^ The inherently heterogeneous patient profiles of HFpEF provide an ideal use case for multi-dimensional phenotypic representation, which we define as *phenomapping*.^17–19^ In the RCT, the Treatment of Preserved Cardiac Function Heart Failure with an Aldosterone Antagonist Trial (TOPCAT), spironolactone did not significantly lower the risk of major adverse cardiovascular events.^20^ Subsequent analyses of TOPCAT have shown heterogeneous treatment effects across participants, requiring an evaluation of the extent to which the RCT cohort is representative of real-world patients.^20–22^

Identifying the complex phenotypic profile of patients with HFpEF in a real-world cohort can quantify the generalizability of TOPCAT across patient populations. We leverage the population of HFpEF patients captured in the electronic health record (EHR) at 5 hospital sites in a large, geographically dispersed health system to demonstrate a strategy to define the representativeness of RCT for both patient characteristics and anticipated real-world treatment effects.

## METHODS

### Data Availability

The TOPCAT cohort is publicly available through the National Heart, Lung, and Blood Institute Biologic Specimen and Data Repository Information Coordinating Center (BioLINCC) The TOPCAT dataset is available at https://biolincc.nhlbi.nih.gov/studies/topcat/. Due to the nature of patient confidentiality, the Yale New Haven Health cohorts are not available. Code for study analysis will be made publicly available at time of publication.

### Study Populations

The first study population, the TOPCAT trial, was a multi-center international RCT that enrolled patients between 2006 and 2012 and evaluated the effect of spironolactone compared with placebo on the incidence of the combined cardiovascular outcome of death from cardiovascular cause, myocardial infarction, stroke, aborted cardiac arrest, and hospitalization for decompensated heart failure among patients with HFpEF. Details of TOPCAT (ClinicalTrials.gov identifier: NCT00094302) have been previously published.^20^ The study enrolled 3445 individuals ≥ 50 years from North America (the United States and Canada), South America (Brazil and Argentina), and Eastern Europe (the Republic of Georgia and Russia) with left ventricular ejection fraction (LVEF) ≥ 45%, one sign and one symptom of heart failure, and at least one hospitalization for heart failure in the preceding 12 months. Alternatively, those without a hospitalization but with an elevated B-type natriuretic peptide were also included. Given the established heterogeneity of the heart failure phenotype in the Eastern European population of TOPCAT, our main analysis included TOPCAT participants from the United States (TOPCAT-US), with sensitivity analyses assessing from TOPCAT participants from the other countries.^21^

The second data source was the Yale New Haven Health System, a large health system that includes several hospitals and associated primary care locations with diverse racial and socioeconomic demographics across Connecticut and Rhode Island. We focused on patients admitted with heart failure to one of the Connecticut sites between January 2015 through April 2023. The study included patients across 5 sites within 4 geographically distinct hospitals: Yale New Haven Hospital York Street Campus (YNHH YSC) and Yale New Haven Hospital St. Raphael’s Campus (YNHH SRC), Greenwich Hospital (GH), Bridgeport Hospital (BH), and Lawrence + Memorial Hospital (LMH). Of note, YNHH YSC and YNHH SRC are located in New Haven, while the other sites are located in other cities/towns in Connecticut. The health system uses an Epic EHR system with data organized in Epic Clarity^®^, a SQL database management system. Strengthening the Reporting of Observational Studies in Epidemiology (STROBE) reporting guidelines were followed. The Yale Institutional Review Board reviewed this study, and a waiver of consent was granted because it was a retrospective study of medical records.

### Outcome

The primary outcome of the TOPCAT RCT was composite cardiovascular mortality including from myocardial infarction, stroke and heart failure.

### EHR Heart Failure Cohort Derivation

Mapping an RCT to an EHR cohort study requires the identification of a cohort that best emulates the eligibility criteria of the study. We extracted patients with at least one hospital admission with a heart failure International Classification of Diseases, Tenth Revision, Clinical Modification (ICD-10-CM) code, representing codes with the root I50. We curated patient encounters with an echocardiogram within six months of the hospitalization and an LVEF of 45% or above and without any prior echocardiogram with an LVEF below 45%. The cohort was then filtered for TOPCAT exclusion criteria (Figure S1). Hospital sites included YNHH YSC, YNHH SRC, BH, GH, and LMH, referred to collectively as the EHR cohorts.

### Curation and Mapping of RCT variables to EHR cohorts

We extracted baseline demographics, conditions, procedures, vital signs, medications, laboratory values, and echocardiogram variables from participant-level data for TOPCAT. We selected structured TOPCAT variables, including continuous variables with <30% missingness in the EHR patients, therefore balancing conditions that were captured in a majority of participants with patient profile completeness. We excluded covariates that provided redundant information or did not have a corresponding definition in the EHR and variables that were related to the trial logistics or its timeline (Table S1). In all, 63 covariates, 1 treatment arm indicator, and 1 outcome of time to composite cardiovascular mortality were included (Figures 1a and 1b, Table S2-S3).

**Figure 1:**
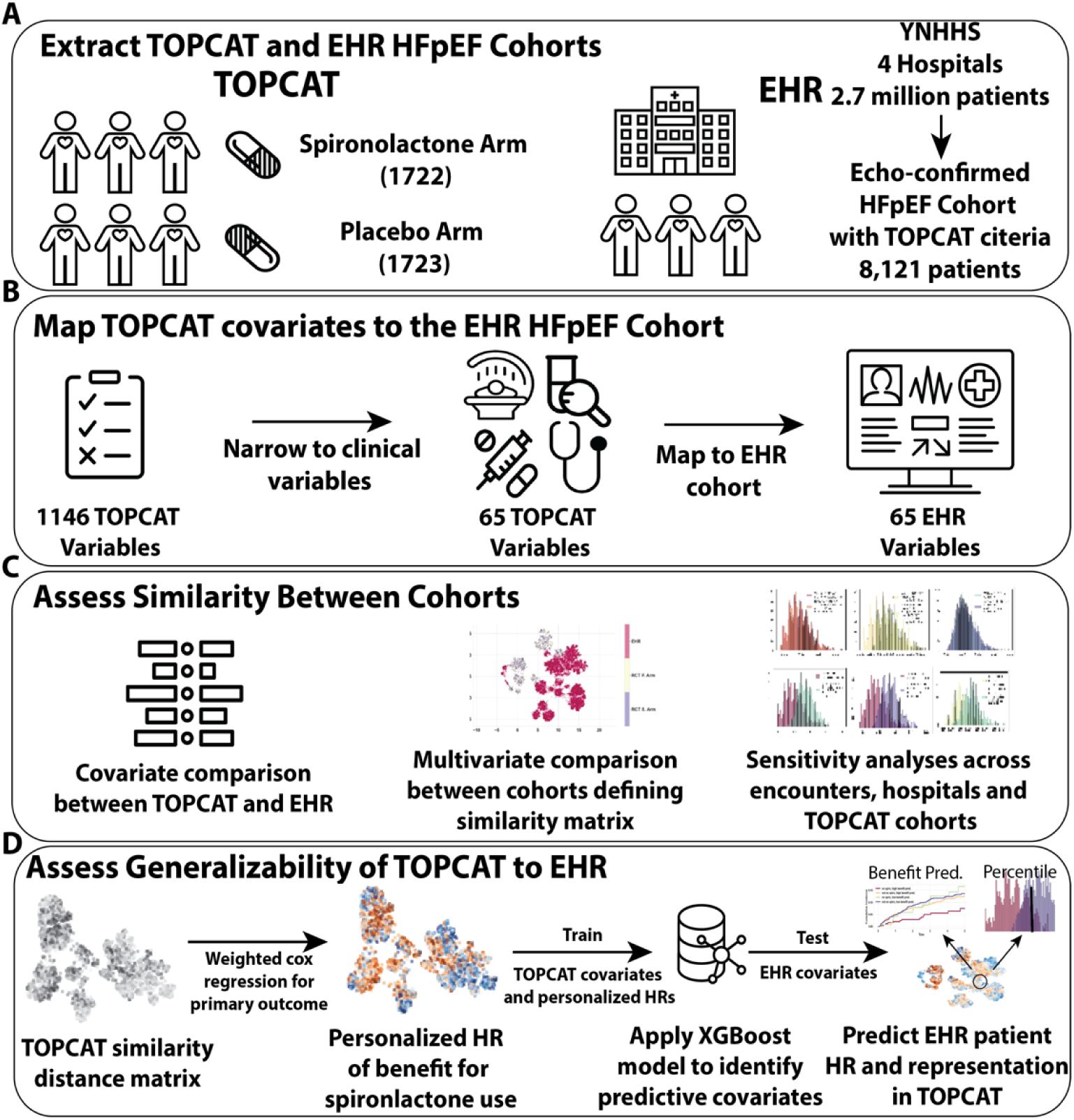
Study Overview. **A:** Depiction of the extraction of TOPCAT and EHR patients with HFpEF. **B:** Narrowing and acquisition of TOPCAT variables to 65 clinically relevant variables with rule-based mapping to EHR variables. **C:** Univariate and multivariate covariate comparison between TOPCAT and EHR patients with HFpEF across multiple sensitivity analyses comparing the different TOPCAT treatment arms and those with and without prior HF hospitalizations with first and last admission encounters within the EHR patients with HFpEF, and two definitions of EHR patients with HFpEF derived by EF. **D:** Assessing the generalizability of TOPAT by deriving personalized hazard ratios for spironolactone use in the TOPCAT participants, training an extreme gradient boosting classifier on the most important covariates for determining these HRs and applying it to the EHR patients with HFpEF, comparing composite cardiovascular outcomes in the EHR patients with HFpEF stratified by predicted personalized benefit and being on spironolactone, and finally assessing the representation of EHR patients with HFpEF in the TOPCAT population by calculating the percentile distance each EHR patient is from the median TOPCAT participant. **Abbreviations:** EHR: Electronic Health Record, HR: Hazard Ratio, YNHHS: Yale New Haven Hospital System

Two clinicians (PT and EKO) collaboratively defined the computational phenotype of each of the 65 TOPCAT variables and outcomes to be deployed in the EPIC Clarity^®^ extracts to map the TOPCAT variables. These included tables summarizing structured data such as conditions, procedures, laboratory values, and medications and semi-structured data such as echocardiogram reports from the EHR. For those with multiple hospitalizations, a random hospitalization encounter was chosen as the start date, and outpatient medications, procedures, conditions, vital signs, and laboratory values were included, if available, to simulate a baseline phenotypic profile similar to the TOPCAT participants (Supplemental Methods).

Each data category required a separate rule-based mapping function with variable ICD-10-CDM code or variable name string-search (Supplemental Methods, Figure 1b, Table S2-S3). EHR patients who died within 30 days of index hospitalization were removed from outcome analysis in order to further match patients with lower acuity to the TOPCAT participants.

TOPCAT and EHR cohorts were pre-processed separately with continuous and binary categorical values. Pre-processing included imputation of missing values, removal of variables with high collinearity, and winsorization of outliers, which followed our previously described methods.^19^ In a sensitivity analysis, the results from two imputation methods, MissForest and multivariate imputation of chained equations MICE, were compared to assess whether the imputation methods, which both use chains of random forests to impute each variable at a time, produced different results.^23,24^ MICE has been used in past EHR imputation studies, but imputation methods to address missingness not at random, as is often the cause in the EHR (Supplemental Methods), are lacking.^25,26^

### Phenotypic Distance Metric to Evaluate Representation Distance Between Cohorts

We defined a metric to summarize the differences across a cohort’s complex multivariate differences by calculating a dissimilarity distance across covariates. We combined distances across the covariate landscape using Gower’s distance, which is a similarity metric that incorporates mixed-type (categorical and continuous) data (Supplemental Methods).^27^ We weighted each covariate distance by its prognostic significance, defined by the beta coefficient of a univariate Cox proportional hazards model to predict the hazard of the combined cardiovascular outcome in the TOPCAT control arm. We refer to this weighted Gower’s distance as the “*phenotypic distance*”. Larger cohorts were subsampled to have the same number of individuals as smaller cohorts for comparison.

To quantitatively assess the distance between cohorts, we compared the median Gower’s distance distribution within a cohort with the distribution between two cohorts. The Gower’s distance, however, is a dimensionless scaled metric of relative distance, so defining this distance between the TOPCAT and EHR cohorts is not directly interpretable. In addition, phenotypic differences exist within each cohort simply due to the variation of phenotypic profiles. To address these issues, we defined the ratio of median phenotypic distances between a cohort’s individuals and a reference cohort’s individuals and between individuals within the reference cohort, named the *phenotypic distance metric (PDM)*, which quantifies the indexed dissimilarity when comparing individuals between two cohorts. A ratio greater than 1 represented a larger difference between cohorts than within one cohort.

In sensitivity analyses, we evaluated median differences between subpopulations of TOPCAT and the five hospital cohorts comparing the median distances within and between subgroups, including the spironolactone and placebo arms of TOPCAT-United States (US), TOPCAT-Non-US, TOPCAT-Americas, and TOPCAT-Eastern-Europe (EE), and the 5 EHR based cohorts.

We also assessed the contribution of changes in population and covariate distributions to the PDM by simulating various distributions and calculating the resulting PDM.

### Individual EHR Patient Representation in RCT

We defined the position of each EHR patient within the phenotypic distribution of TOPCAT patients across prognostically relevant covariates. For this, we recalculated the weighted Gower’s distance of each EHR cohort patient from the TOPCAT-US cohort covariates most prognostic for the combined cardiovascular outcome. Next, we defined an index TOPCAT-US participant, the one most phenotypically representative of TOPCAT-US participants, based on the shortest phenotypic distance to all other TOPCAT-US participants. We estimated the representation of each EHR cohort patient in TOPCAT-US by calculating their percentile of phenotypic distance from the index TOPCAT-US participant. Specifically, using the TOPCAT-US cohort distribution of phenotypic distance from the index TOPCAT-US participant, we determined the percentile of each EHR cohort patient within this distribution, representing the position of each EHR patient within the phenotypic distribution of TOPCAT-US patients.

### RCT-derived clinical effect estimates against treatment patterns in the EHR

Based on our prior work^19,28^, we also used the results from TOPCAT to define which patients from the EHR would benefit from spironolactone treatment as a function of patient covariates (Supplemental Methods). We first identified neighborhoods within two separate TOPCAT sub-populations (TOPCAT-US and TOPCAT-EE) with high Gower similarities between participants. We then estimated individualized hazard ratios for the primary TOPCAT outcome for the neighborhoods of high-similarity participants. We then built XGBoost models based on the baseline characteristics and outcomes of the TOPCAT-US participants to identify patients in the EHR predicted to benefit from spironolactone use. We assessed spironolactone use in the clinical setting by EHR patients with an expected benefit from spironolactone (iHR<1). To compare, we trained a second XGBoost model on baseline characteristics and outcomes of TOPCAT-EE participants and compared the proportion of EHR patients estimated to benefit from spironolactone.

### Statistical Analysis

We summarized categorical variables by number and proportion present in each group and continuous variables as mean and standard deviation or median and interquartile range. Categorical variables were compared between the two cohorts using a chi-squared test and continuous variables using Welch’s two-sided t-test.^29^ We also calculated the absolute standardized mean difference for each covariate between each cohort pair and the median standardized mean difference with IQR for each TOPCAT-EHR cohort pair and each pair of EHR cohorts.^29^

We calculated the PDM as described above with interquartile values of the metric. We depicted the qualitative difference between the TOPCAT cohort and the EHR cohort by projecting the phenotypic distances onto a dimensionality reduction method called uniform manifold approximation and projection (UMAP).^19,30,31^ The method projects the high-dimensional dataset onto two dimensions by ensuring points are closest to their nearest neighbors while also attempting to preserve the global representation of each point in the manifold.

The Yale Institutional Review Board reviewed this study, and a waiver of consent was granted because it was a retrospective study of medical records.

## RESULTS

### Populations Characteristics

The US participants within the TOPCAT trial included 1151 participants with a median age of 71 (63-80, 25-75% IQR) years and included 557 (48%) women. Of the trial population, 572 (50%) were assigned to the spironolactone arm and 579 (50%) to the placebo control arm. The EHR cohorts included 30,858 patients with a diagnosis of heart failure. Of these, 12,548 (41%) had one or more hospitalizations with a principal or a secondary diagnosis of heart failure (91,404 hospitalizations overall) and had at least one echocardiogram across either inpatient or outpatient settings, demonstrating an LVEF ≥45% and no prior echocardiograms with an LVEF <45%. There were 11,712 patients who had at least one echocardiogram within six months of their hospital admission, similar to the TOPCAT inclusion criteria. When filtering by the TOPCAT exclusion criteria, 8,121 patients remained in the final cohort, 7,738 having a recorded outcome at least 30 days after entering the cohort (Figure S1). Among the index hospitalization randomly chosen for each patient, 2526 (31%) patients were treated at YNHH YSC, 2482 (30%) at YNHH SRC, 1654 (20%) at BH, 403 (5%) at GH, and 1056 (13%) at LMH. Across these sites, the median age of patients ranged from 77 (IQR 68-86) years to 85 (IQR 77-91) years. The proportion of women ranged from 54% to 62%. Overall, the EHR cohorts were older and had a higher proportion of women compared with TOPCAT-US, and in addition a higher proportion of minorities compared with TOPCAT-Non-US (Table 1, Table S4).

**Table 1:**
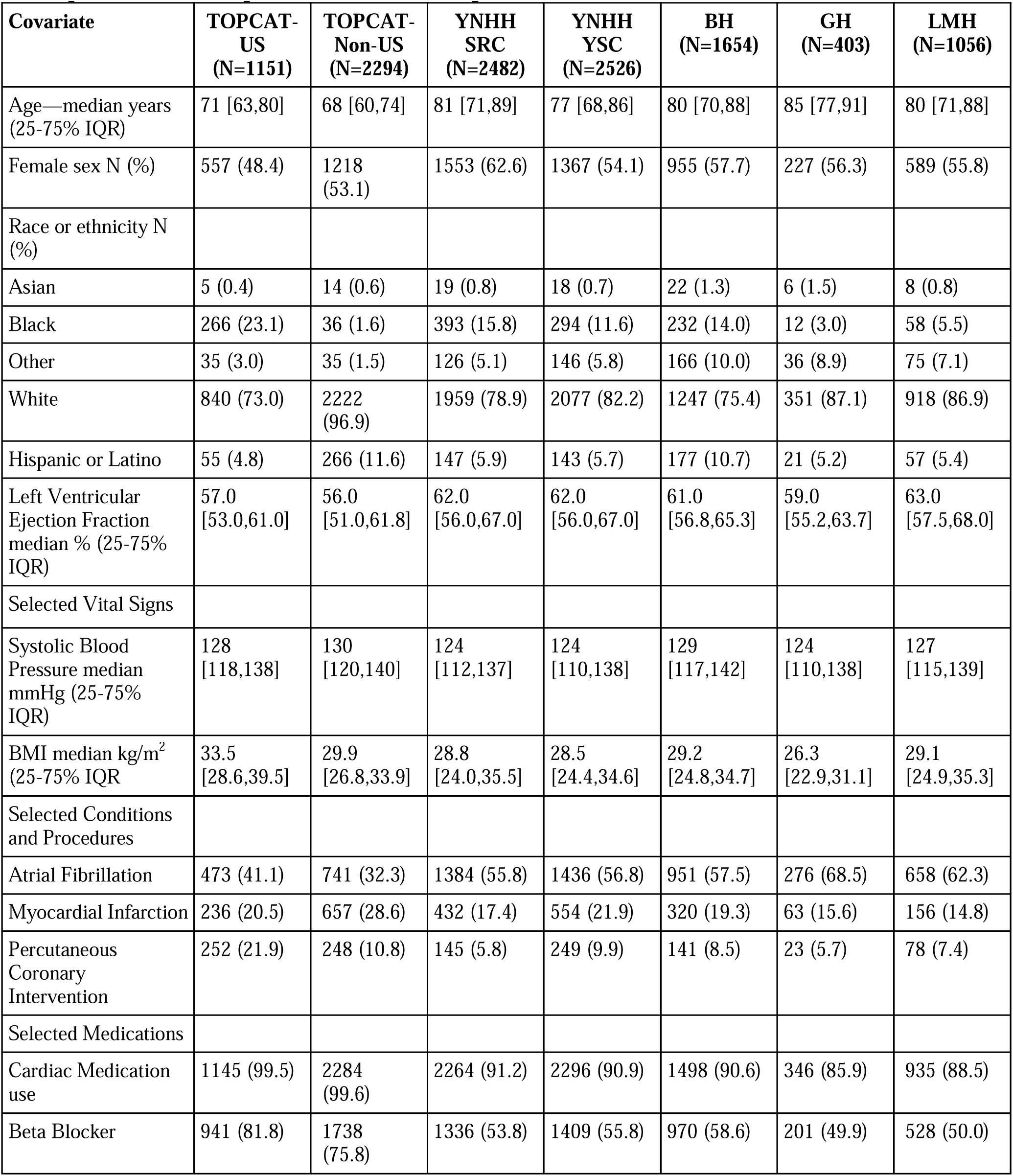
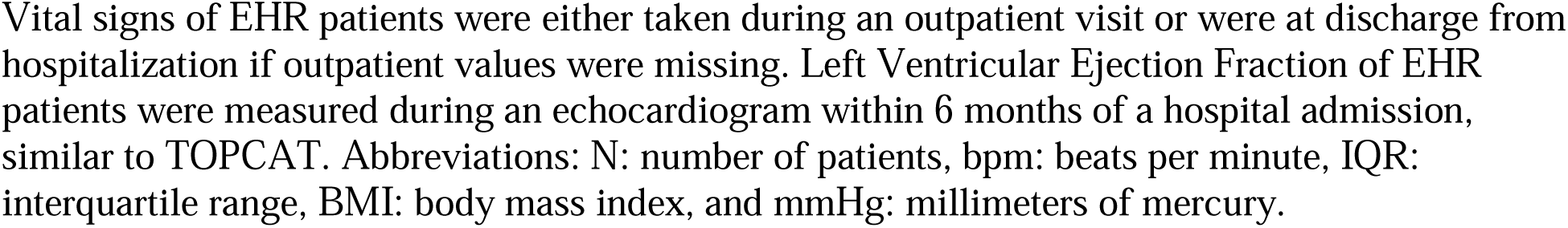
Selected Population Characteristics of the TOPCAT participants compared with EHR patients with HFpEF in 5 different hospital sites.

### Similarity and representation between RCT and real-world EHR cohorts

The median absolute standardized mean difference (ASMD) across baseline covariates between the EHR patients and TOPCAT-US participants was higher than within the EHR patients themselves (median ASMD 0.09, IQR 0.03-0.17 vs median ASMD 0.26, IQR 0.10-0.52) (Table S5). Both by qualitative UMAP visualizations (Figure S2-S3) and quantitative similarity distance comparisons (Table 2), there was less phenotypic separation between the TOPCAT-US and EHR cohorts (Figure 2a and 2c, S4-S7) than between the TOPCAT-EE and EHR cohorts (Figures 2b, Figures S8-S9) as evidenced by a PDM of 1.03 (IQR 0.96-1.09 vs 1.45 (IQR 1.26-1.62).Further, we confirmed the phenotypic similarity between the treatment and placebo arms of TOPCAT defined by a PDM of 1.00 (IQR 0.99-1.00) and estimated a median PDM between the EHR of 1.00 (IQR 0.98-1.02). In an additional sensitivity analysis, we found the PDM between pooled EHR cohorts and the non-US TOPCAT population to be 1.30 (IQR 1.15-1.48) (Table 2, Figure S10).

**Figure 2:**
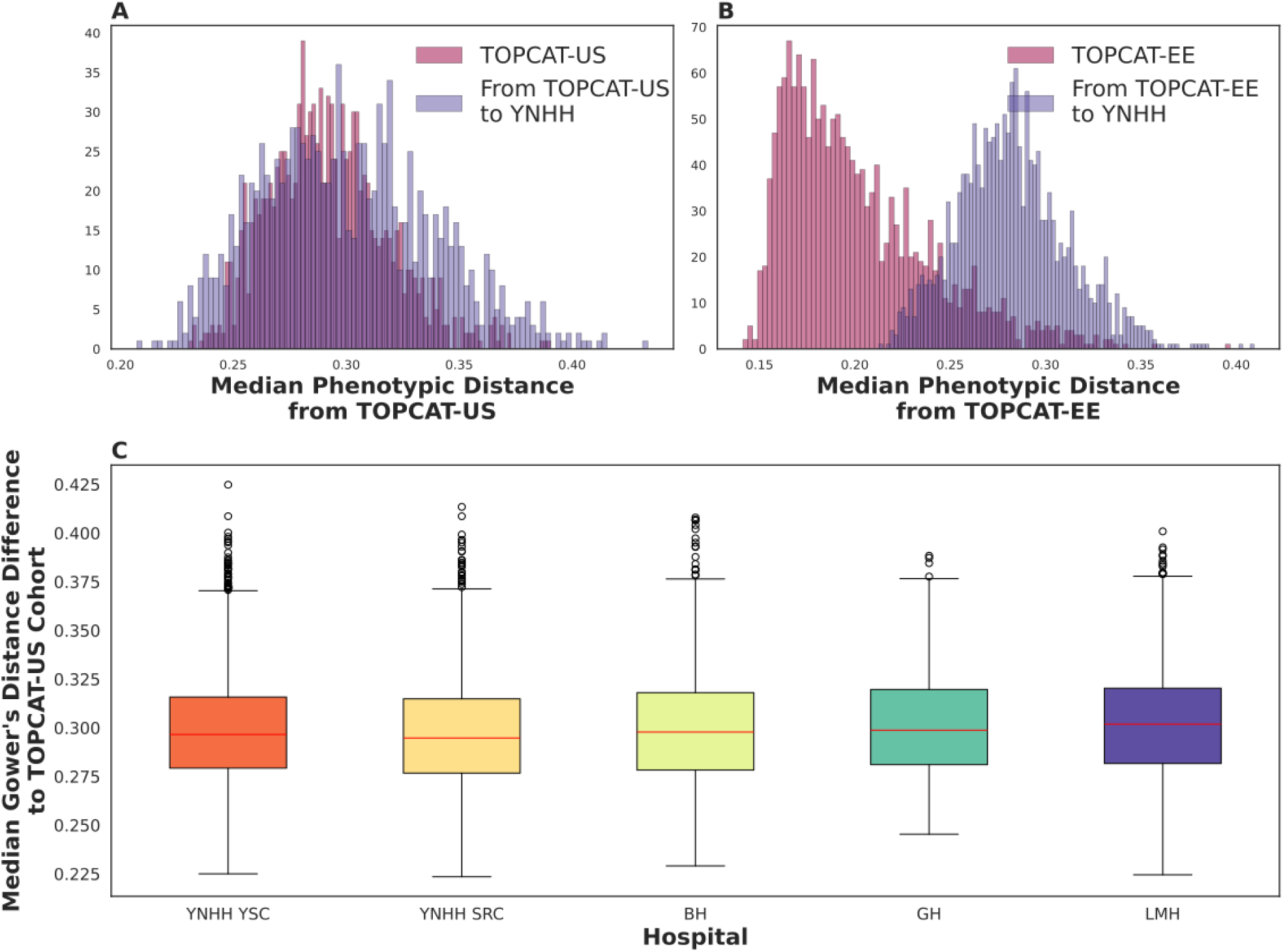
Median Phenotypic Distance between TOPCAT subgroups and EHR patients. **A:** Histogram of median phenotypic distance between TOPCAT-US (Dark red) and the EHR cohort (Purple). **B:** Histogram of median phenotypic distance between the TOPCAT-Eastern Europe (EE) cohort (Dark red) and EHR cohort (Purple). **C:** Median phenotypic distance difference of each subgroups from the entire EHR cohort median phenotypic distance from TOPCAT-US. Orange is York Street Campus (YNHH YSC), Yellow is St. Raphael’s Campus (SRC), Lime green is Bridgeport Hospital (BH), Green in Greenwich Hospital (GH), and purple is Lawrence and Memorial Hospital (LMH).

**Table 2:**
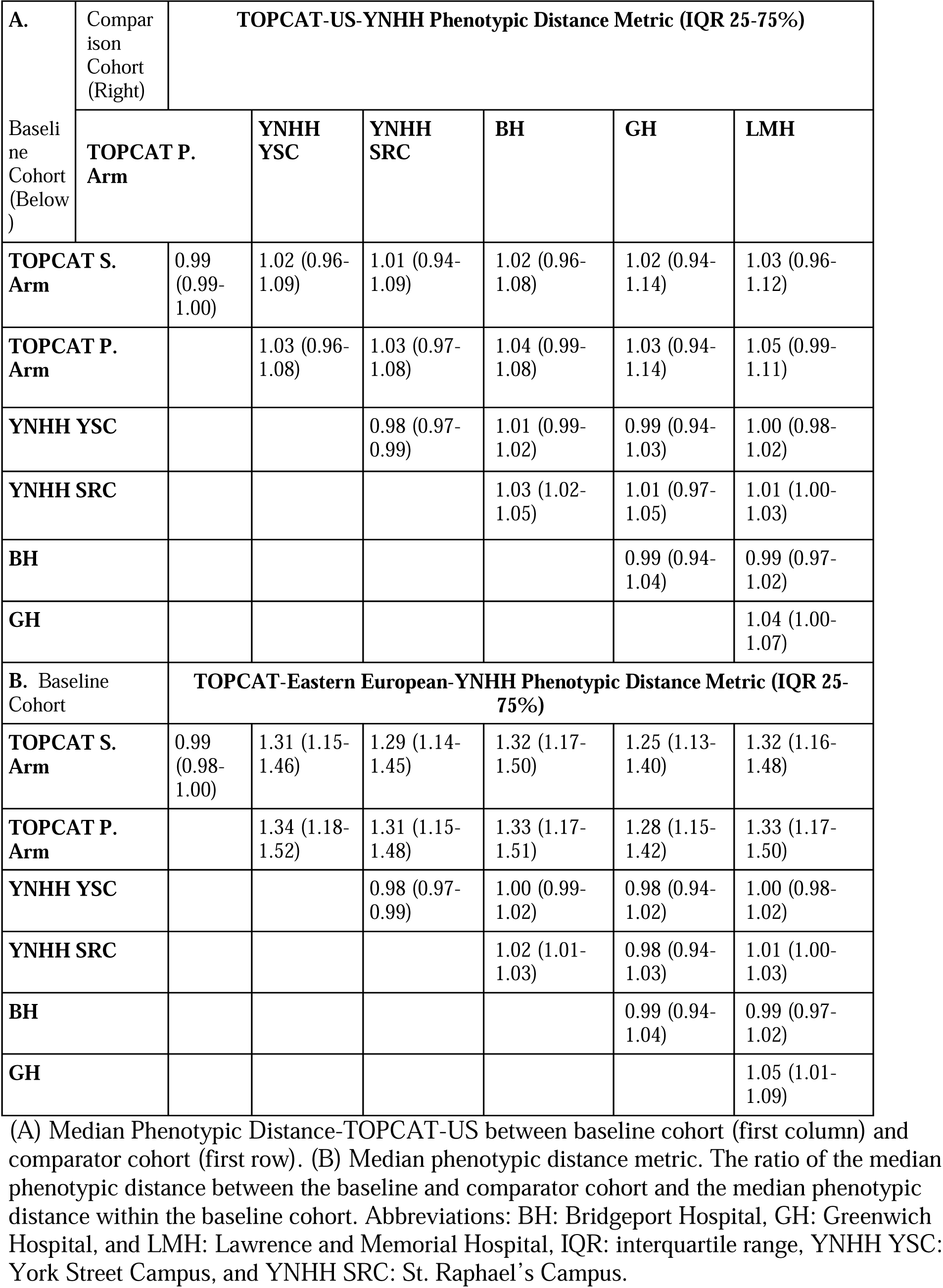
Median Phenotypic Distance Metric Across Cohorts.

When comparing imputation methods, across 12 phenotypic distributions stratified by individual distance pair (TOPCAT-TOPCAT, TOPCAT-EHR, and EHR-EHR) and by TOPCAT-subpopulation (TOPCAT-US, TOPCAT-Non-US, TOPCAT-Americas, and TOPCAT-Eastern-Europe), there were largely no differences in phenotypic distance distribution between MICE and MF imputation (Figure S10). With regards to real-world individual representation within TOPCAT, we compared all EHR patients to the most representative TOPCAT participant, or the individual closest in phenotypic distance to all other participants, termed the index TOPCAT participant. All patients in the EHR cohorts were further in phenotypic distance from the index TOPCAT participant than 47% of the TOPCAT participants. In addition, the average patient in the EHR cohorts was further from the index TOPCAT patient than 83% of the TOPCAT patients.

In a simulation assessing the effect of varying population distributions, we found the population distance distribution does influence PDMs varying between 1.04 and 6.01(Figure S11). In a simulation assessing the effect of varying covariate value distributions, we found varying the covariate distribution changed the PDM by 1-3% for 26/36 (continuous covariate) and 13/18 (categorical covariate) simulations (Figures S12-20).

### Outcomes in real-world cohorts based on expected therapeutic effects in the RCT

Within TOPCAT, 671 patients experienced the primary outcome, 362 of which were from TOPCAT-US. Based on the individualized treatment effects (ITE) model developed in TOPCAT-US participants, all (7,738) EHR patients with a recorded outcome within the assessed time period were predicted to benefit from spironolactone use (iHR of <1). Of these, 1,208 (16%) were receiving the medication before outcome measurement (Figure 3a, Table S6). In contrast, from the ITE model derived from TOPCAT-Eastern European participants, 13% of EHR patients were predicted to benefit from spironolactone use (Figure 3b, Table S7).

**Figure 3:**
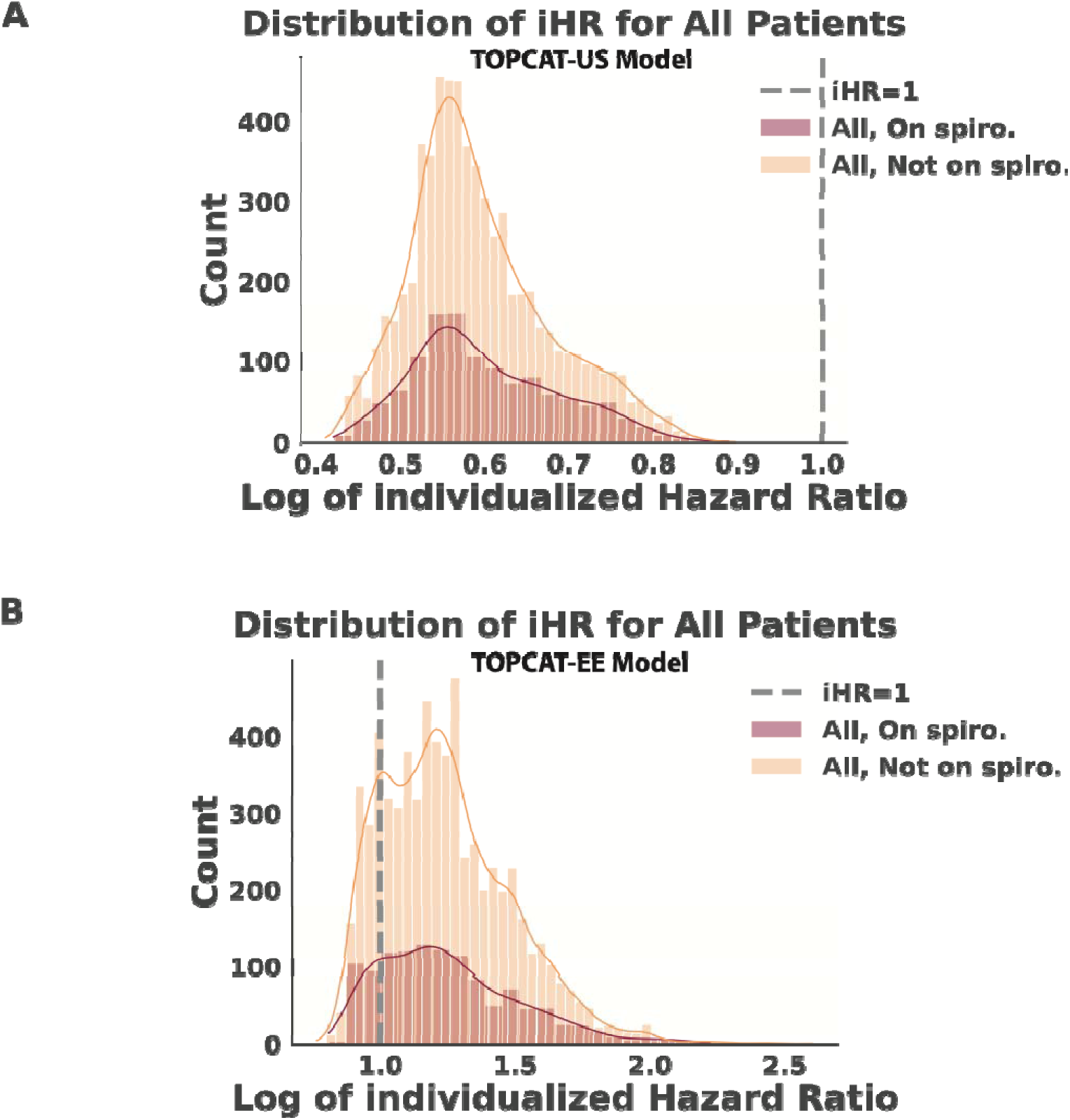
Distribution of individualized hazard ratios of EHR cohort and Kaplan-Meier curve of EHR patients with high predicted spironolactone benefit. **A:** Distribution of log of individualized hazard ratios of time to cardiovascular event stratified by spironolactone use predicted for the EHR outcome cohort from the TOPCAT-US participants. The vertical line represents an individualized hazard ratio of 1, and values to the left of the dotted line represent high predicted benefit of spironolactone use. Red represents patients on spironolactone, orange represents patients not on spironolactone. **B:** Distribution of log of individualized hazard ratios of time to cardiovascular event stratified by spironolactone use predicted for the EHR outcome cohort from the TOPCAT-Eastern Europe participants. The vertical line represents an individualized hazard ratio of 1, and values to the left of the dotted line represent high predicted benefit of spironolactone use. Red represents patients on spironolactone, orange represents patients not on spironolactone. **Abbreviations:** EE: Eastern Europe, Spiro.: spironolactone, iHR: individualized hazard ratio for time to cardiovascular event stratified by spironolactone use.

## DISCUSSION

In this study, we demonstrate a strategy to quantify the multi-dimensional representation gap between an RCT and real-world patients in the EHR by presenting an approach to map computable phenotypes of RCT participants to real-world clinical populations. We propose a quantitative metric that computes information across all available covariate axes to define a unifying similarity score across cohorts, and apply the score with individualized outcome information in the RCT to identify EHR patients most likely to benefit from the RCT intervention. The PDM quantified TOPCAT participants from the US to be more similar to the EHR patients compared with TOPCAT participants from Eastern Europe, which is expected given the known controversy of the proportion of TOPCAT patients from Eastern Europe having heart failure.^21^ In addition, all individual EHR patients were further in phenotypic distance from a representative TOPCAT-US participant compared with the 47% of TOPCAT-US participants, supporting quantification of the representation gap. Finally, the TOPCAT-US participants, who historically had been found to have a significant mortality benefit from spironolactone, predicted all EHR patients would likely have a positive response to spironolactone use, while the TOPCAT-Eastern-Europe participants only predicted 13% of EHR patients to benefit. This confirms that there is greater representation of the EHR patients in TOPCAT-US participants compared with TOPCAT-Eastern-Europe.

Our study builds upon the literature evaluating the representativeness of RCTs for real-world settings. Prior approaches have focused on defining differences across a limited number of covariate axes and have also largely been used on one-to-one comparisons across singular covariate or solely categorical data, with the information across covariate comparisons interpreted qualitatively.^12–15,32–34^ Two complementary methods to PDM measure population differences between RCTs and real-world patient populations, but with key differences in scope and methodology. The GIST 2.0 metric, for example, measures the weighted proportion of an EHR population that follows an RCT’s eligibility criteria.^14^ This is valuable for identifying what proportion of a patient population would be eligible for an RCT, but it does not compare baseline clinical characteristics between cohorts at the individual level.^14^ The Trial score provides an estimate of how close an individual is from the average TOPCAT patient using 9 continuous variables weight by beta coefficients from a logistic regression predicting composite cardiovascular outcome.^33^ Our approach complements this by assessing similarity weighted by outcome across each TOPCAT-EHR pair, taking into account the distribution of distances between all individuals. By quantifying the differences between cohorts across multiple axes, we overcome the fallacy of comparing the average distribution of covariates across the entire population against the averages across another population. This ignores stark differences in various clinical subpopulations that are not identified by focusing on the average, as evidenced by the differing PDMs found between TOPCAT subpopulations and the EHR patients. In addition, we applied a quantitative phenotypic distant metric across five sites within four different hospitals that demonstrates the flexibility of the application across different cohorts.

Our study has important clinical implications. When clinicians assess whether a patient would benefit from a particular treatment, they often refer to relevant RCTs for intervention in medical practice. They must also, however, look at the patient in front of them and determine how effectively the study translates to care. They ascertain (1) how generalizable is the study result to my patient? and (2) was my patient well-represented in this trial? The clinician considers not only demographics or comorbidities but also the entire picture of the patient. Our approach addresses this decision quantitatively and by providing the phenotypic distance metric to assess where the patient in question lies along the phenotypic distribution of the RCT participants and to suggest whether the patient may benefit from the intervention based on phenotypic similarity to the RCT participants. Our approach to generalize ITEs from an RCT to a real-world population predicts the response to the RCT-tested intervention in the new population guided by phenotypic similarity.

Our study uses TOPCAT as an example of RCT to demonstrate a process to capture the complex multidimensional picture of each RCT participant and to compare them directly to real-world patients such as those in the EHR. We describe a method to predict benefits in the real-world population based on individual patient characteristics and covariates deemed important by RCT individuals and also assess their representation in the RCT. Our quantification of the large representation gap between TOPCAT and EHR patients with HFpEF suggest an overarching need to assess representation from a multi-dimensional perspective during trial implementation and interim analysis.^35^ This further supports an increased role of tracking trial recruitment against real-world populations. In addition, the degree of external validity of a new population can be quantified by the PDM.

Another observation from our work is the intensive nature of mapping data from clinical trials to real-world populations in the EHR. We highlight the challenges and variability of translating RCT cohorts and study variables to the EHR setting, suggesting another impedance to translating RCT evidence to our patients. Common data models such as the Observational Medical Outcomes Partnership standardize EHR mapping to make this research easier to accomplish and apply across multinational institutions.^36^ As trials increasingly become pragmatic, there is an urgent need to computably define RCT conditions within the context of the EHR since the manual approach to identify key features will represent a challenge for trial operations.

There are limitations that merit consideration. Representation of real-world populations in the EHR is a unique challenge since the data represent a snapshot of time when the patient interacts with the medical system, and the patients seeking care likely represent a subset of the HFpEF population. The patients included, however, represented five sites with unique and diverse patient populations, thus maximizing the possible landscape of patients with HFpEF. Second, we chose a set of covariates available across the RCT and the EHR, which may not be a fully representative set of conditions that differ between individuals. However, features captured in large RCTs are often comprehensive, and our clinician-led approach designed a strategy to map many of these conditions using text phrases, billing codes, and all possible patient encounters in the system. We also confirmed the robustness of our approach with sensitivity analyses that focused on prognostically relevant conditions with similar results. Third, we chose Gower’s distance given its flexibility with modeling both categorical and continuous data and its ability to weigh some conditions more relative to the others. Although this represents one of many Euclidean distances appropriate for assessing differences across covariates, it has been found superior to the other methods in identifying phenotypic differences.^37^ Finally, given the inherent biases in prescribing interventions and treatment in the EHR such as ascertainment bias and confounding by indication, the PDM does not purport to predict the composite outcome of a new patient population--rather it assesses which patients of the new patient population are most similar to the original RCT, and therefore are likely to experience outcome as what was seen in the trial.

We propose a novel approach to evaluating the real-world representativeness of RCT participants against corresponding patients in the EHR across the full multidimensional spectrum of the represented phenotypes. This enables the evaluation of the implications of RCTs for real-world patients.

## Data Availability

The TOPCAT cohort is publicly available through the National Heart, Lung, and Blood Institute Biologic Specimen and Data Repository Information Coordinating Center (BioLINCC) The TOPCAT dataset is available at https://biolincc.nhlbi.nih.gov/studies/topcat/. The Yale electronic health record cohorts are not available due to the use of patient data.

## DISCLOSURES

Dr. Thangaraj, Dr. Oikonomou, and Dr. Khera are coinventors of a provisional patent not related to the current work (63/606,203). Dr. Khera is an Associate Editor of JAMA and receives research support, through Yale, from the Blavatnik Foundation, Bristol-Myers Squibb, Novo Nordisk, and BridgeBio. He is a coinventor of U.S. Provisional Patent Applications 63/177,117, 63/428,569, 63/346,610, 63/484,426, 63/508,315, 63/580,137, 63/562,335, and a co-founder of Ensight-AI, Inc and Evidence2Health, LLC. Dr. Oikonomou is an academic co-founder of Evidence2Health LLC, and has been a consultant for Caristo Diagnostics, Ltd and Ensight-AI, Inc. He is a co-inventor in patent applications (US17/720,068, 63/619,241, 63/177,117, 63/580,137, 63/606,203, 63/562,335,WO2018078395A1, WO2020058713A1) and has received royalty fees from technology licensed through the University of Oxford. Dr. Suchard receives grants and contracts from the US Food & Drug Administration, the US Department of Veterans Affairs and Johnson & Johnson, all outside the scope of this work.

## FUNDING

The study is supported by the National Heart, Lung, and Blood Institute of the National Institutes of Health (R01HL167858 and K23HL153775). Dr. Thangaraj, Dr. Oikonomou, and Mr. Jayaram are also supported by grants from the National Institutes of Health (5T32HL155000-03, 1F32HL170592-01, and T35HL007649 respectively).

## SUPPLEMENTAL MATERIALS

Supplemental Methods

Supplemental Results

Figures S1-S24

Tables S1-S9

References 38-40

## Notes

### Funding Statement

The study is supported by the National Heart, Lung, and Blood Institute of the National Institutes of Health (R01HL167858). Dr. Thangaraj and Dr. Oikonomou are also supported by grants from the National Institutes of Health (5T32HL155000-03 and 1F32HL170592-01, respectively).

### Summary of Updates

We have updated all sections to include more sensitivity analyses, methodology comparisons, and implications discussions.

