## Supplementary Information for "Computational Phenomapping of Randomized Clinical Trials to Enable Assessment of their Real-world Representativeness and Personalized Inference"

| Title | Page |
|---|---|
| **Supplementary Methods** | 3-8 |
| **Supplementary Results** | 9 |
| **Figure S1: Flowchart for Final EHR HFpEF cohort when considering TOPCAT exclusion criteria.** | 10 |
| **Figure S2: UMAP visualization of TOPCAT-US-EHR phenotypic overlap with phenotypic distance as metric.** | 11 |
| **Figure S3: UMAP visualization of EHR patient overlap separated by site.** | 12 |
| **Figure S4: Median Phenotypic Distance across cohorts TOPCAT-US, YNHH YSC, YNHH SRC, BH, GH, and LMH.** | 13 |
| **Figure S5: Median Phenotypic Distance across cohorts TOPCAT-US, YNHH YSC, YNHH SRC, and BH.** | 14 |
| **Figure S6: Median Phenotypic Distance across cohorts TOPCAT-US, YNHH YSC, GH, and LMH.** | 15 |
| **Figure S7: Median Phenotypic Distance across cohorts YNHH SRC, BH, GH, LMH.** | 16 |
| **Figure S8: Median Phenotypic Distance across cohorts TOPCAT-Eastern-European, YNHH YSC, YNHH SRC, and BH.** | 17 |
| **Figure S9: Median Phenotypic Distance across cohorts TOPCAT-Eastern-European, GH, and LMH** | 18 |
| **Figure S10: The Distribution of Phenotypic Distance Metrics.** | 19 |
| **Figure S11: Simulation assessing the influence of different TOPCAT-EHR phenotypic distance distributions with the same mean on the PDM** | 20 |
| **Figure S12: Simulated White Blood Cell Count and Statin Use covariate distributions with calculated Phenotypic Distance Metric.** | 21 |
| **Figure S13: Simulated Blood Urea Nitrogen Level and Atrial Fibrillation covariate distributions with calculated Phenotypic Distance Metric.** | 22 |
| **Figure S14: Simulated Blood Urea Nitrogen Level and Statin Use covariate distributions with calculated Phenotypic Distance Metric.** | 23 |
| **Figure S15: Simulated Systolic Blood Pressure and Atrial Fibrillation covariate distributions with calculated Phenotypic Distance Metric.** | 24 |
| **Figure S16: Simulated Systolic Blood Pressure and Percutaneous Coronary Intervention covariate distributions with calculated Phenotypic Distance Metric.** | 25 |
| **Figure S17: Simulated Systolic Blood Pressure and Statin Use covariate distributions with calculated Phenotypic Distance Metric.** | 26 |

| | |
|---|---|
| **Figure S18: Simulated White Blood Cell Count and Atrial Fibrillation covariate distributions with calculated Phenotypic Distance Metric.** | 27 |
| **Figure S19: Simulated White Blood Cell Count and Percutaneous Coronary Intervention covariate distributions with calculated Phenotypic Distance Metric.** | 28 |
| **Figure S20: Simulated Blood Urea Nitrogen Level and Percutaneous Coronary Intervention** | 29 |
| **Figure S21: Features Ranked by importance using XGBoost Model trained on TOPCAT-US participants.** | 30 |
| **Figure S22: SHAP analysis train on TOPCAT-US participants showing feature importance for prediction of personalized benefit for spironolactone use.** | 31 |
| **Figure S23: Features Ranked by importance using XGBoost Model trained on TOPCAT- Eastern-European participants.** | 32 |
| **Figure S24: SHAP analysis train on TOPCAT-Eastern-European participants showing feature importance for prediction of personalized benefit for spironolactone use.** | 33 |
| **Table S1: Examples of TOPCAT trial variables removed from the study analysis organized by category.** | 34 |
| **Table S2: Mapping of TOPCAT covariates to EHR definitions.** | 35-36 |
| **Table S3: Mapping of TOPCAT covariate medication definitions to EHR patient datasets.** | 37 |
| **Table S4: Comprehensive Baseline Characteristics of TOPCAT participants and EHR patients used in this study analysis.** | 38-42 |
| **Table S5: Median Standardized Mean Difference of each covariate for pairs of TOPCAT participants and EHR patients used in this study analysis.** | 43-47 |
| **Table S6: Incidence Rates of EHR Patients with Predicted benefit by TOPCAT-US participants from Spironolactone.** | 48 |
| **Table S7: Incidence Rates of EHR Patients with Predicted benefit by TOPCAT-Eastern Europe participants from Spironolactone.** | 49 |
| **Table S8: Missingness of continuous variables in TOPCAT and the EHR patient datasets.** | 51 |
| **Table S9: Missingness of categorical variables in TOPCAT trial** | 52 |
| **SUPPLEMENTAL REFERENCES** | 53-54 |

### Supplementary Methods

#### General covariate category encounter extraction rules

Conditions and cardiac procedures were derived from structured information in the medical history list, current problem list, and the ICD-10-CM diagnosis codes and procedure names recorded across all encounter types. Medications were preferentially defined by outpatient medication lists, but inpatient medication administration data were used if outpatient medication data were not available.

#### Rule Based Mapping of TOPCAT covariates to EHR variables

1. Demographics were pulled from the hospital encounter, which included age, sex, and race/ethnicity. Race was divided into four categories based on the TOPCAT variables: "Asian", "Black", "White", or "other," and ethnicity was either "Hispanic/Latino" or none. Patients with multiple races and ethnicities were included in multiple categories. Race descriptors in the EHR from East Asian and South Asian countries were considered "Asian". Middle eastern, Native American, and Pacific Islander race descriptors were included in the "Other" category.
2. Conditions defined in TOPCAT variables were mapped by ICD10 code and diagnosis name extracted from Epic Clarity. This was done by a text search of both the corresponding ICD-10-CM code and corresponding diagnosis name (Table S2). The ICD-10-CM codes chosen included all relevant diagnosis names, which were manually evaluated by a clinician. We maximized the number of condition events found for each patient by combing multiple tables, including hospital and outpatient diagnosis encounters, medical history, and problem list. All conditions occurring before and during the index hospital encounter were included.

3. Procedures defined in TOPCAT variables were mapped in a similar process to conditions, except for an additional text search of procedure names were used from a procedure encounter list in addition to ICD-10-CM diagnosis codes for presence of the cardiac device or procedure. All procedure encounters occurring before the hospital encounter were included. We further identified additional encounters of cardiac device implantation and procedures, including defibrillators, pacemakers, coronary artery bypass grafting (CABG) and percutaneous coronary interventions (PCI) in the procedure hospital encounters table. To identify applicable procedure character sequences, we manually evaluated all unique procedure names to identify those corresponding to each procedure. For example, the string "pace" produced only names corresponding to a pacemaker and the string "defib" produced only names corresponding to defibrillator (Table S2).
4. Laboratory measurements were extracted based off the "base" laboratory name available (Table S2). The most recent outpatient lab value at least one week from the hospitalization encounter. If no outpatient value was available, the most recent resulted laboratory value for each laboratory measurement in each hospital encounter was used. This sequence served to minimize missingness and use a laboratory value closest to discharge from the hospital, which would be most like the TOPCAT participants.
5. TOPCAT medication variables were mostly grouped by drug class, except for some variables that were combinations of other TOPCAT variables. We manually mapped all possible generic medications corresponding to each relevant drug class or TOPCAT medication variable (Table S3). The medication name or character sequence was identified by the simple generic name and within a table extracting hospital medication administration. We also extracted from an outpatient medication list. Included outpatient medications started no later

than one week after discharge from the hospital encounter and discontinued no earlier than one week before admission to the hospital encounter.

6. Vital signs of heart rate, systolic and diastolic blood pressure, weight, and height were extracted from outpatient encounter flow sheets at least one week from the index hospitalization. If an outpatient value was not available, the value closest to hospitalization discharge was used to most closely resemble the TOPCAT participants. BMI was calculated from weight and height.
7. Outpatient medications, procedures, and conditions more than a week before the hospital admission were included. The most recent outpatient vital signs and laboratory values more than a week from hospital admission were included in order to best estimate baseline laboratory and vital sign values. If these values were missing in outpatient, values were used at the time of discharge of the hospital encounter.
8. Across all 11,712 patients of the EHR cohorts, 7,086 patients had similarity and representation between RCT and real-world EHR cohorts' outpatient laboratory values and vital signs, 8,935 patients had outpatient laboratory values, 9,633 had outpatient vital signs, and 575 had only inpatient laboratory values and vital signs.

**Harmonization and Imputation of TOPCAT and EHR dataset**

Age, EF, vital signs, and lab measurements were all considered continuous variables, while all other variables were binary categorical. All continuous variable values were converted to numeric float values, and all binary variables were converted to categorical values. Missing binary variables were considered absent and assigned a value of 0. Missing continuous values and negative values were considered missing and assigned "NaN" in preparation for imputation. Any "Yes" or "checked" values were assigned "1" and "No", "0", "Female", "1", and "Male",

“0”. Any lab values with “<” or “>” were rewritten to the boundary value (i.e., “>6000” was 6000) and commas were removed.

We compared the imputation of missing values using MissForest and multivariate imputation of chained equations (MICE). We imputed with MissForest by running the *MissForest* package from *missingpy* in python, which chains random forests to predict imputed values that minimize the root mean square between complete and imputed data. This algorithm has been shown to be the most effective imputation algorithm in mixed-type categorical and continuous data and high-dimensional data.[23,24] We utilized the *IterativeImputer* class of *SciKit Learn* in python to impute missing variables with MICE, which models missing values from each variable as a function of the other variables.[26] One lab value, hematocrit, and two medication categories, anti-hypertensive medications and non-cardiovascular medications, were dropped due to >0.9 collinearity with other variables. Outliers were removed by winsorizing to the 2.5% and 97.5% percentiles.

**Gower’s Distance calculation**

Continuous variable distances were calculated as the absolute value difference divided by range, and binary variable distance was assigned 1 if identical and 0 otherwise. The distance per patient was then calculated as the average across all variables.[27,31]

**UMAP Projection Display Parameters**

UMAP used the calculated weighted Gower values as the “precomputed” distance metrics. Number of neighbors was 10, minimum distance was 0.5, number of components was 2.

**Development of TOPCAT phenomap and calculation of individualized treatment effects (ITE)**

In order to estimate ITE in the TOPCAT RCT, we applied the Trialmap method developed by Oikonomou et al.[19,28,31] Briefly, Gower's distances between all TOPCAT participants were calculated. The participants were weighted by a kernel based method, specifically:

$$(1 - Gower's\ distance)^{15}$$

Anything below the median weight was set to zero. A cox proportional hazards model for composite cardiovascular outcome with individuals weighted by the kernal method was built and an individualized hazard ratio (iHR) for the outcome for each participant was estimated. The ITEs were calculated separately for each TOPCAT subpopulation, such as TOPCAT-US and TOPCAT-EE.

**Generalization of TOPCAT ITE results to EHR patients**

Using an application of the Trialmap method[19,28,31], An XGBoost machine learning was trained on all TOPCAT baseline characteristics and treatment group to predict individualized hazard ratios for each participant. Separate models were created for two TOPCAT subpopulations (i.e. TOPCAT-US and TOPCAT-EE). The XGBoost model was optimized using an 80/20 training/validation split. The most important features were then selected using the Borutashap method, which combines Boruta and SHAP (SHapely Additive exPlanations) principles.[38] Specifically, Boruta builds shadow features which are permutations of the covariates from the training set with correlations removed.[39] Using the SHAP feature importance algorithm, any features more important than the shadow feature is considered important. The SHAP feature importance algorithm determines the contribution of each covariate to the outcome for each individual.[40] After selecting the most important features, the model was optimized for root mean squared error using the hyperparameter tuning function *RandomizedSearchCV* (Parameters: “eta” or learning rate: [0.01, 0.05, 0.10, 0.15], “max_depth'”or maximum depth of each tree: [3, 5, 6, 10, 15, 20],  “subsample” or proportion of training samples used: [0.5, 1.0, 0.1], 'colsample_bytree' or proportion of features used for each trea: [0.4, 1.0, 0.1], 'colsample_bylevel' or proportion of features used for each level of the tree : [0.4, 1.0, 0.1], and 'n_estimators' or number of trees used: [100, 500, 1000]). After the model was hypertuned, the final model was applied to the EHR cohort to predict individualized hazard ratios for each patient.

**Computing**

All coding, data analysis, and most plots were completed in Python. Python packages used included pandas, numpy, scipy, os, collections, matplotlib, gower, tableone, umap, scikitlearn, and lifelines.

### Supplementary Results

#### TOPCAT variable mapping to EHR variables missingness.

Out of the 1143 variables available in the TOPCAT RCT data dictionary, 65 (5.8%) variables were able to be mapped to EHR data. Within the EHR data, labs were missing in 805 (7%) to 7843 (67%) of patients, with the median missingness of labs being in 2766 (24%) of patients. All other continuous variables (vital signs, Age, ejection fraction) were missing in 0-297 (0-3%) of patients (Table S8). Within the TOPCAT cohort, 16 (76%) of continuous variables had 0-1% missingness, and the rest had 3-46% missingness.

#### BorutaSHAP analysis results

BorutaSHAP analysis of the TOPCAT-US iHR predictive model found six covariates that showed significant feature importance: whether a patient had atrial fibrillation, whether they were on a statin, their systolic blood pressure, and their laboratory values of blood urea nitrogen, fasting glucose, and white blood cell count (Figure S21-S22). The TOPCAT-Eastern Europe (EE) iHR predictive model found eight covariates with significant feature importance including the gender of the patient, whether they had diabetes mellitus or dyslipidemia, whether they were on a statin, a nitrate, an “other” anti-hypertension medication, or an “other” cardiovascular medication, and their blood urea nitrogen level (Figure S23-S24).

### Supplementary Figures and Figure Legends

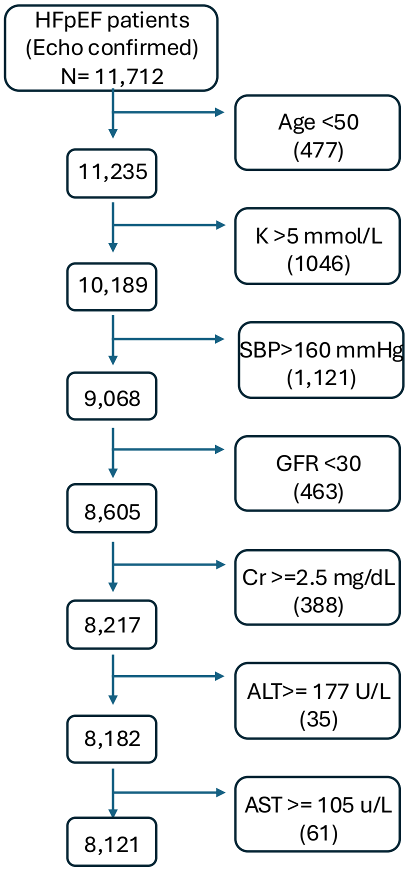

**Figure S1: Flowchart for Final EHR HFpEF cohort when considering TOPCAT exclusion criteria.** Abbreviations: ALT: Alanine Aminoransferase, AST: Aspartate Aminotransferase, Cr: Creatinine, GFR: Glomerular Filtration Rate, dL: deciliter, K: Potassium, L: Liter, mg: milligram, mmHg: milligrams of mercury, mmol: millimolar, SBP: Systolic Blood Pressure, U: Unit

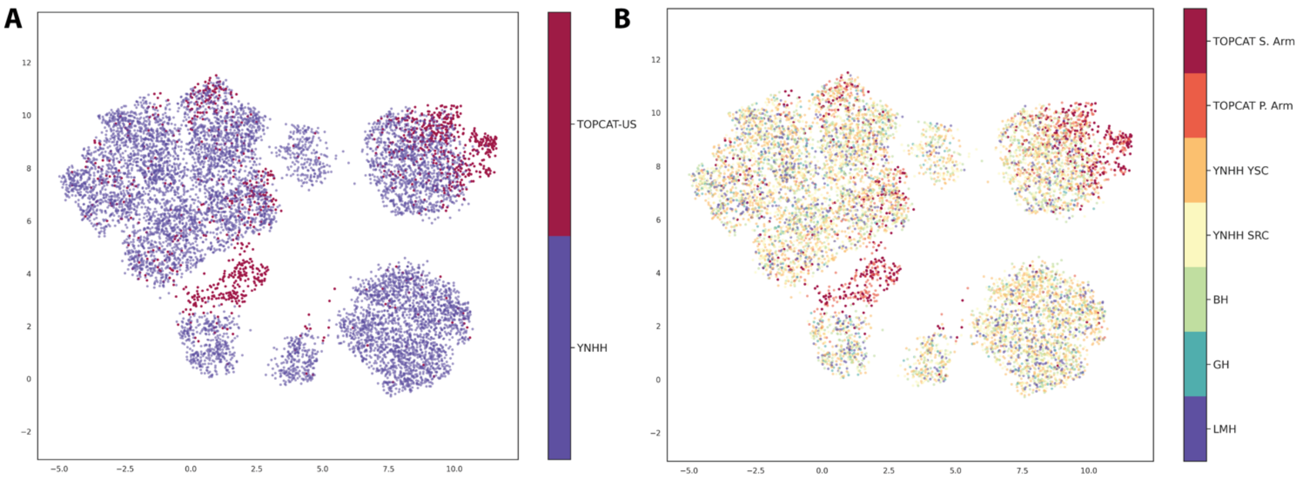

**Figure S2: UMAP visualization of TOPCAT-US-EHR phenotypic overlap with phenotypic distance as metric.** A: TOPCAT-US participants are in Red, YNHHS EHR cohort is in dark purple, B: UMAP-visualization colored by subgroup: Dark red is TOPCAT-US S. Arm, dark orange is TOPCAT-US P. Arm, YNHH YSC is in orange, YNHH SRC is in yellow, BH is in green, GH is in blue, and LMH is in purple. Abbreviations: BH: Bridgeport Hospital, GH: Greenwich Hospital, LMH: Lawrence + Memorial Hospital, P: Placebo, S.: Spironolactone TOPCAT: Treatment of Preserved Cardiac Function Heart Failure with an Aldosterone Antagonist Trial, YNHHS: Yale New Haven Hospital System, YNHH SRC: Yale New Haven Hospital St. Raphael's Campus, YNHH YSC: Yale New Haven Hospital York Street Campus

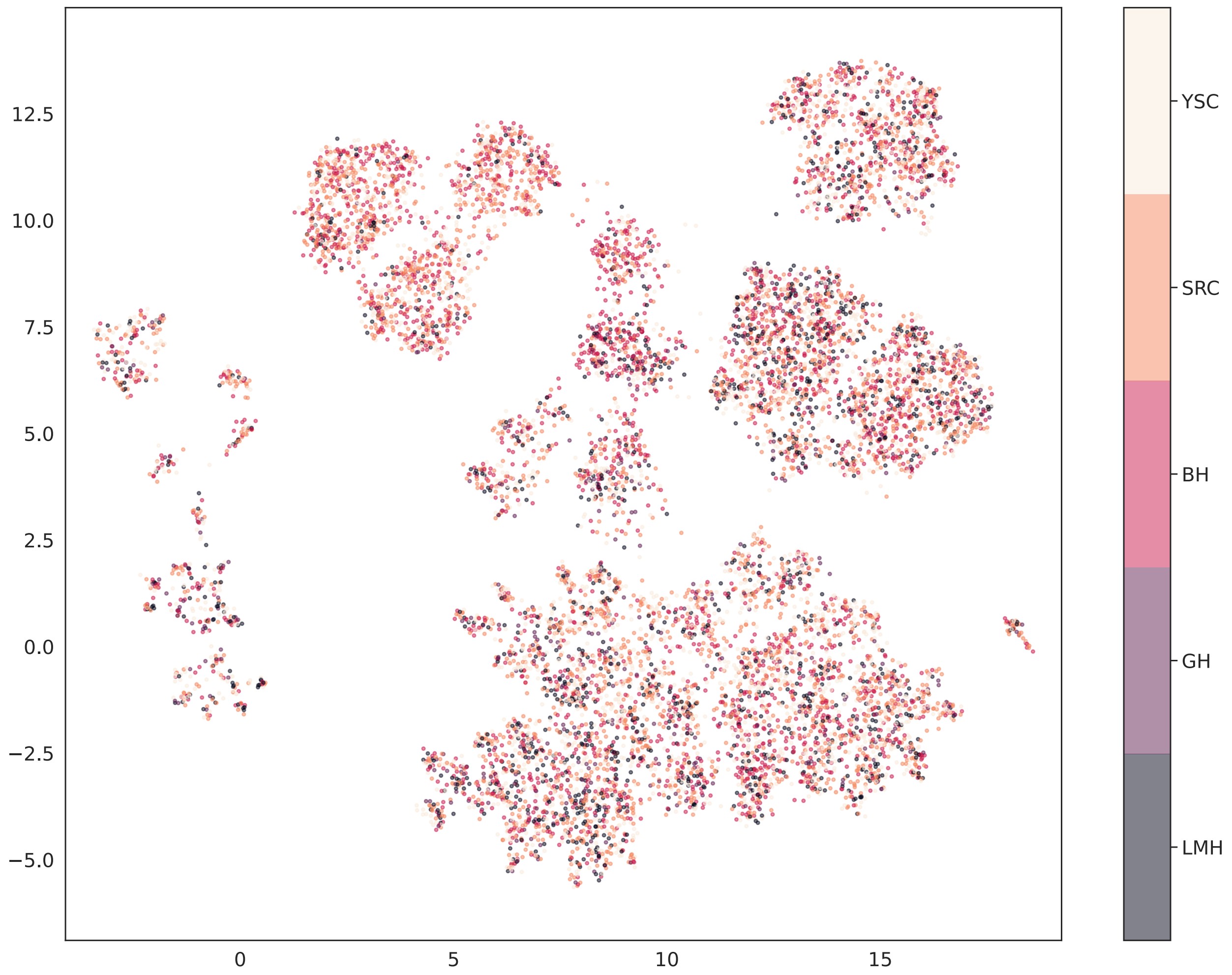

**Figure S3: UMAP visualization of EHR patient overlap separated by site.** Yellow (YSC) is YNHH York Street Campus, Peach (SRC) is YNHH St. Raphael's Campus, Pink (BH) is Bridgeport Hospital, Purple (GH) is Greenwich Hospital, and Dark Grey (LMH) is Lawrence + Memorial Hospital. Abbreviations: BH: Bridgeport Hospital, GH: Greenwich Hospital, LMH: Lawrence and Memorial Hospital, SRC: St. Raphael's Campus, YNHH: Yale New Haven Hospital, YSC: York Street Campus

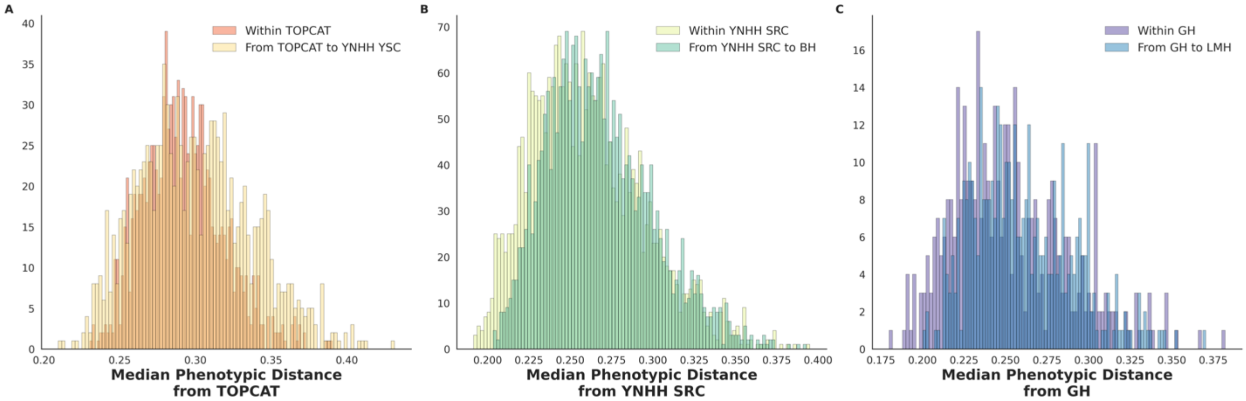

**Figure S4: Median Phenotypic Distance across cohorts TOPCAT-US, YNHH YSC, YNHH SRC, BH, GH, and LMH.** Within each panel, the top labeled histogram represents the median baseline phenotypic distance between participants within (A) Orange, the TOPCAT-US cohort, (B), Light Green, the YNHH SRC cohort, and (C) Purple, the GH cohort. The bottom labeled histogram in each panel represents the median phenotypic distance of the baseline cohort from the comparator cohort:  In (A), Yellow is the median phenotypic distance from the TOPCAT-US cohort to the YNHH YSC cohort, (B) Green is the median phenotypic distance of the YNHH SRC cohort to the BH cohort, (C) Blue is the median phenotypic distance from the GH cohort to LMH cohort. Abbreviations: BH: Bridgeport Hospital, GH: Greenwich Hospital, LMH: Lawrence and Memorial Hospital, TOPCAT: Treatment of Preserved Cardiac Function Heart Failure with an Aldosterone Antagonist Trial, US: United States, YNHH SRC: Yale New Haven Hospital St. Raphael's Campus, YNHH YSC: Yale New Haven Hospital York Street Campus

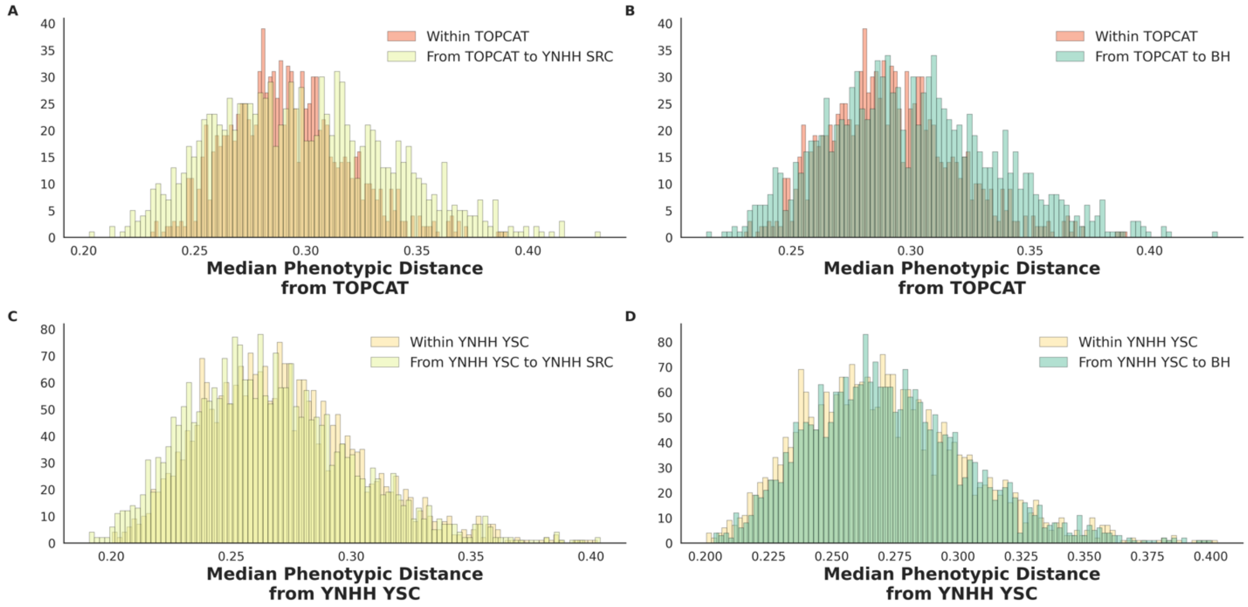

**Figure S5: Median Phenotypic Distance across cohorts TOPCAT-US, YNHH YSC, YNHH SRC, and BH.** Within each panel, the top labeled histogram represents the median baseline phenotypic distance between participants within (A) and (B) Orange, the TOPCAT cohort, and within (C) and (D) Yellow, the YNHH YSC cohort. The bottom labeled histogram in each panel represents the median phenotypic distance of the baseline cohort from the comparator cohort: In (A), Light Green is the median phenotypic distance from the TOPCAT cohort to the YNHH SRC cohort, (B) Green is the median phenotypic distance from the TOPCAT cohort to the BH cohort, (C) Light green is the median phenotypic distance from the YNHH YSC cohort to the YNHH SRC cohort, and (D) Green is the median phenotypic distance from the YNHH YSC cohort to the BH cohort. Abbreviations: BH: Bridgeport Hospital, TOPCAT: Treatment of Preserved Cardiac Function Heart Failure with an Aldosterone Antagonist Trial, US: United States, YNHH SRC: Yale New Haven Hospital St. Raphael’s Campus, YNHH YSC: Yale New Haven Hospital York Street Campus

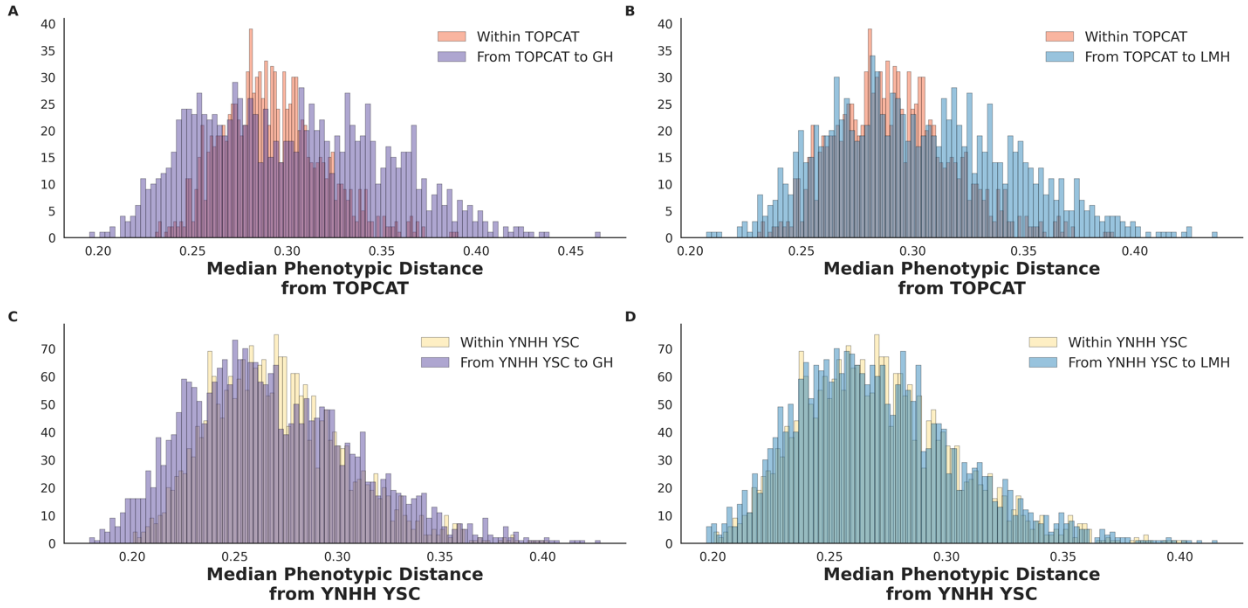

**Figure S6: Median Phenotypic Distance across cohorts TOPCAT-US, YNHH YSC, GH, and LMH.** Within each panel, the top labeled histogram represents the median baseline phenotypic distance between participants within (A) and (B) Orange, the TOPCAT cohort, and within (C) and (D) Yellow, the YNHH YSC cohort. The bottom labeled histogram in each panel represents the median phenotypic distance of the baseline cohort from the comparator cohort: In (A), Purple is the median phenotypic distance from the TOPCAT cohort to the GH cohort, (B) Blue is the median phenotypic distance from the TOPCAT cohort to the LMH cohort, (C) Purple is the median phenotypic distance from the YNHH YSC cohort to the GH cohort, and (D) Blue is the median phenotypic distance from the YNHH YSC cohort to the LMH cohort. Abbreviations: BH: Bridgeport Hospital, GH: Greenwich Hospital, LMH: Lawrence + Memorial Hospital, US: United States, YNHH SRC: Yale New Haven Hospital St. Raphael's Campus.

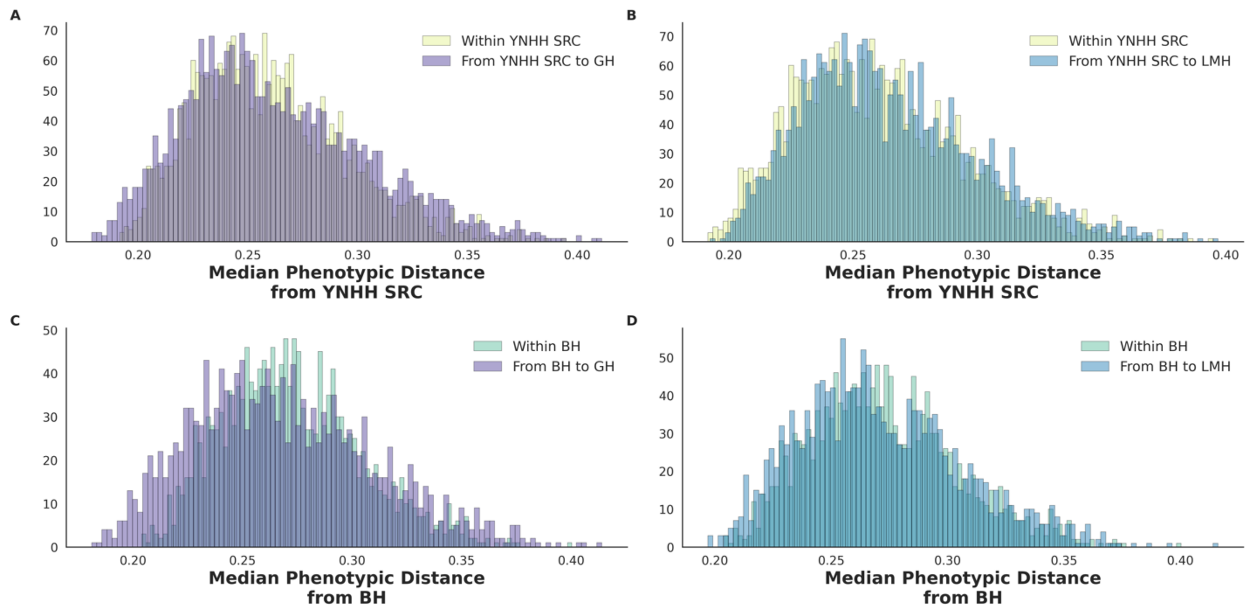

**Figure S7: Median Phenotypic Distance across cohorts YNHH SRC, BH, GH, LMH.** Within each panel, the top labeled histogram represents the median baseline phenotypic distance between participants within (A) and (B) Light green, the YNHH SRC cohort, and within (C) and (D) Green, the BH cohort. The bottom labeled histogram in each panel represents the median phenotypic distance of the baseline cohort from the comparator cohort: In (A), Purple is the median phenotypic distance from the YNHH SRC cohort to the GH cohort, (B) Blue is the median phenotypic distance from the YNHH SRC cohort to the LMH cohort, (C) Purple is the median phenotypic distance from the BH cohort to the GH cohort, and (D) Blue is the median phenotypic distance from the BH cohort to the LMH cohort. Abbreviations: GH: Greenwich Hospital, LMH: Lawrence + Memorial Hospital, TOPCAT: Treatment of Preserved Cardiac Function Heart Failure with an Aldosterone Antagonist Trial, US: United States, YNHH SRC: Yale New Haven Hospital St. Raphael's Campus, YNHH YSC: Yale New Haven Hospital York Street Campus.

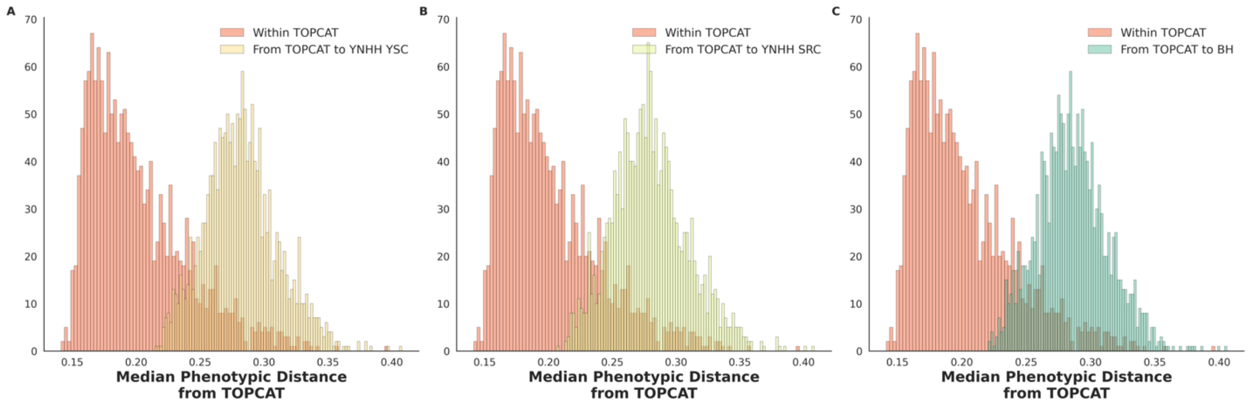

**Figure S8: Median Phenotypic Distance across cohorts TOPCAT-Eastern-European, YNHH YSC, YNHH SRC, and BH.** Within each panel, the top labeled histogram represents the median baseline phenotypic distance between participants within (A-C) Orange, the TOPCAT-Eastern-European cohort. The bottom labeled histogram in each panel represents the median phenotypic distance of the baseline cohort from the comparator cohort: In (A), Yellow is the median phenotypic distance from the TOPCAT-Eastern-European cohort to the YNHH YSC cohort, (B) Light Green is the median phenotypic distance of the TOPCAT-Eastern-European cohort to the YNHH SRC cohort, (C) Green is the median phenotypic distance from the TOPCAT-Eastern-European cohort to BH cohort. Abbreviations: BH: Bridgeport Hospital, GH: Greenwich Hospital, LMH: Lawrence and Memorial Hospital, TOPCAT: Treatment of Preserved Cardiac Function Heart Failure with an Aldosterone Antagonist Trial, YNHH SRC: Yale New Haven Hospital St. Raphael's Campus, YNHH YSC: Yale New Haven Hospital York Street Campus

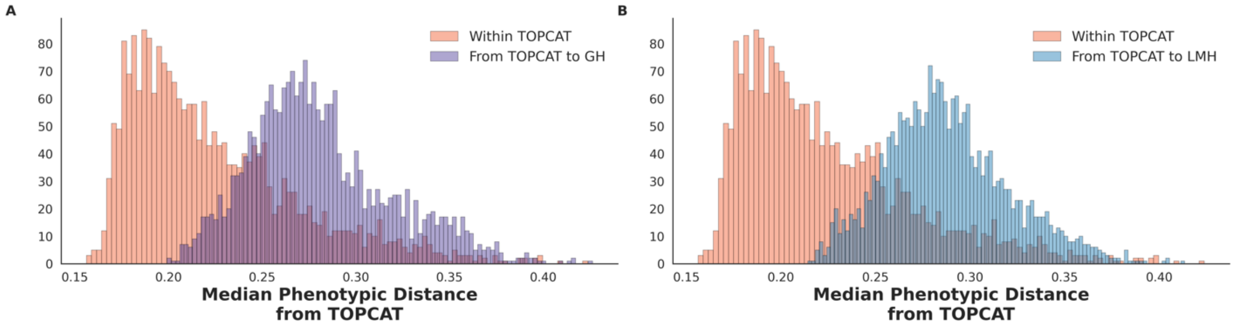

**Figure S9: Median Phenotypic Distance across cohorts TOPCAT-Eastern-European, GH, and LMH.** Within each panel, the top labeled histogram represents the median baseline phenotypic distance between participants within (A-B) Orange, the TOPCAT- Eastern-European cohort. The bottom labeled histogram in each panel represents the median phenotypic distance of the baseline cohort from the comparator cohort: In (A), Purple is the median phenotypic distance from the TOPCAT-Eastern-European cohort to the GH cohort, (B) Blue is the median phenotypic distance of the TOPCAT-Eastern-European cohort to the LMH cohort. Abbreviations: BH: Bridgeport Hospital, GH: Greenwich Hospital, LMH: Lawrence and Memorial Hospital, TOPCAT: Treatment of Preserved Cardiac Function Heart Failure with an Aldosterone Antagonist Trial, YNHH SRC: Yale New Haven Hospital St. Raphael's Campus, YNHH YSC: Yale New Haven Hospital York Street Campus

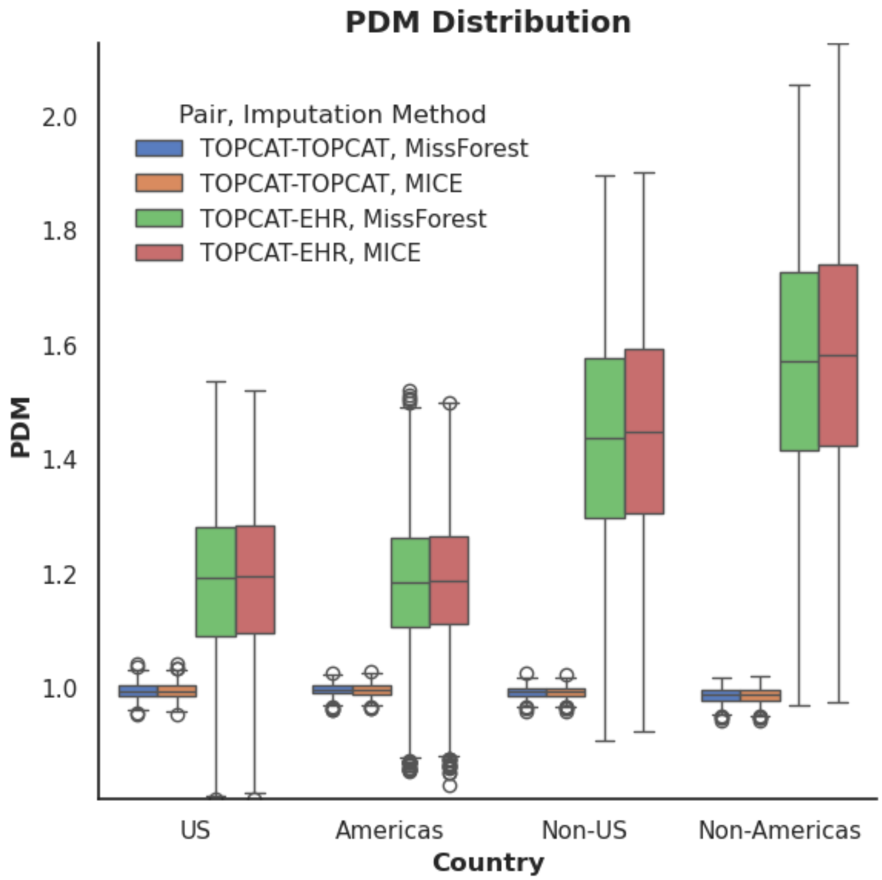

**Figure S10: The Distribution of Phenotypic Distance Metrics** (A) between TOPCAT participants and YNHH EHR participants and (B) between TOPCAT participants stratified by TOPCAT participant location.

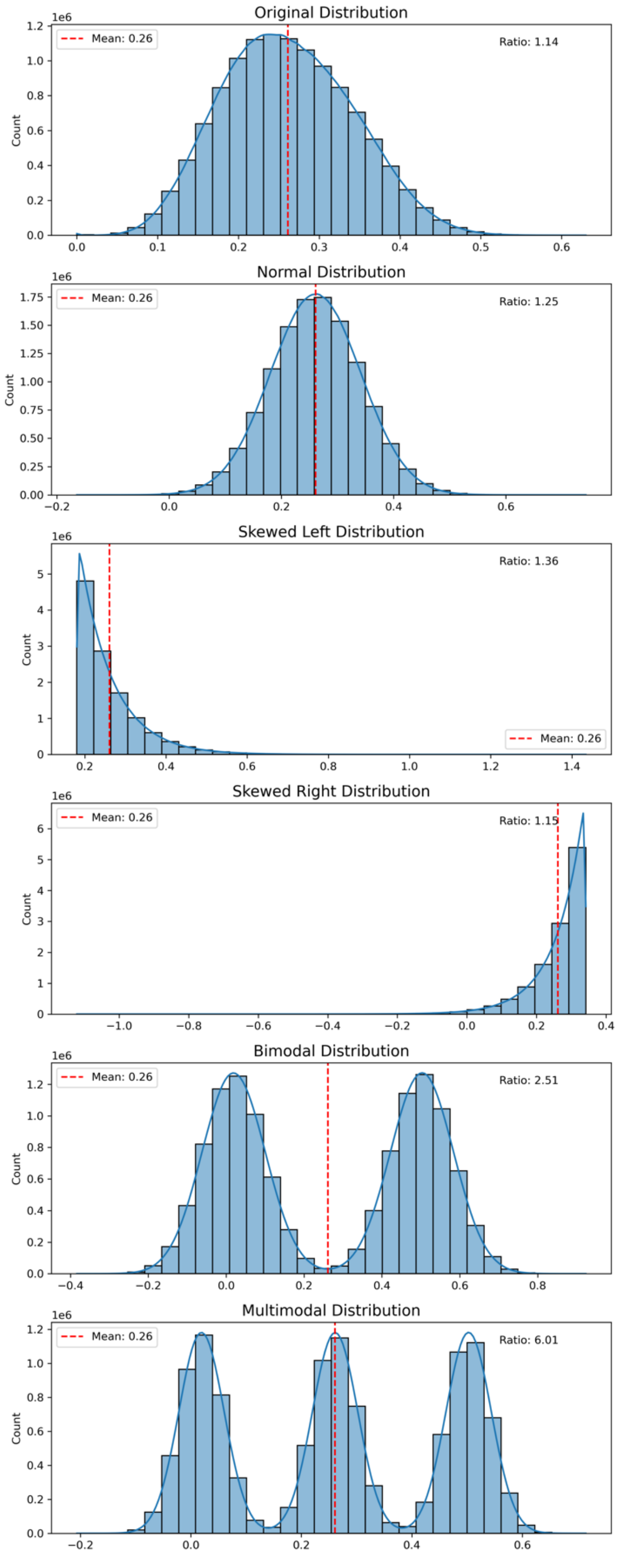

**Figure S11: Simulation assessing the influence of different TOPCAT-EHR phenotypic distance distributions with the same mean on the PDM**

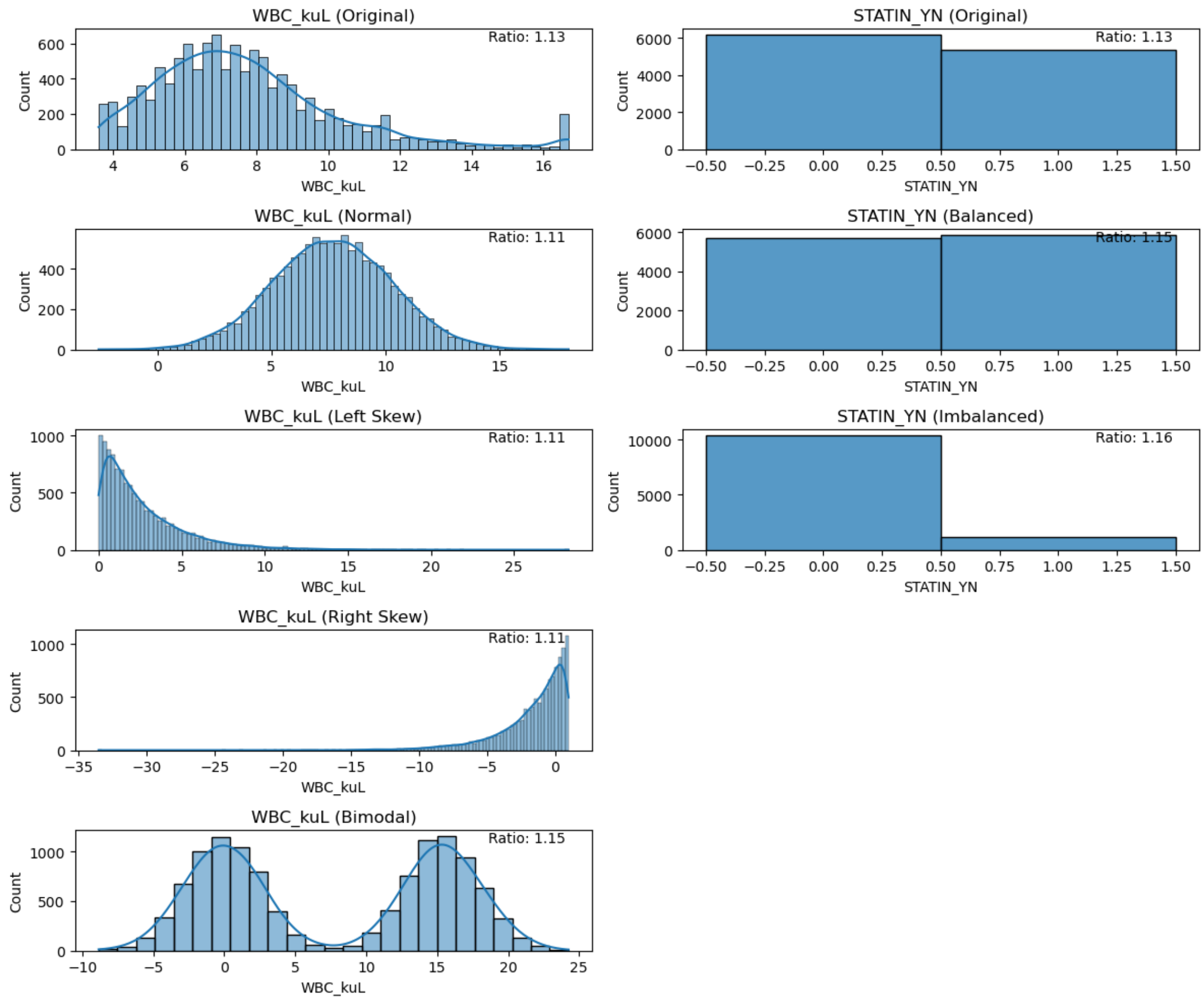

**Figure S12: Simulated White Blood Cell Count and Statin Use covariate distributions with calculated Phenotypic Distance Metric.** Left column depicts the original white blood cell count distribution along with normal, skewed, and bimodal transformations with the Phenotypic Distance Metric (ratio) in the upper right hand corner. Right column depicts the original proportion of patients who use (Right bar) or do not use (Left bar) a statin, balanced proportion (50/50 split) and an imbalanced (90/10 split) proportion with the Phenotypic Distance Metric (ratio) in the upper right hand corner. Abbreviations: WBC: White Blood Count.

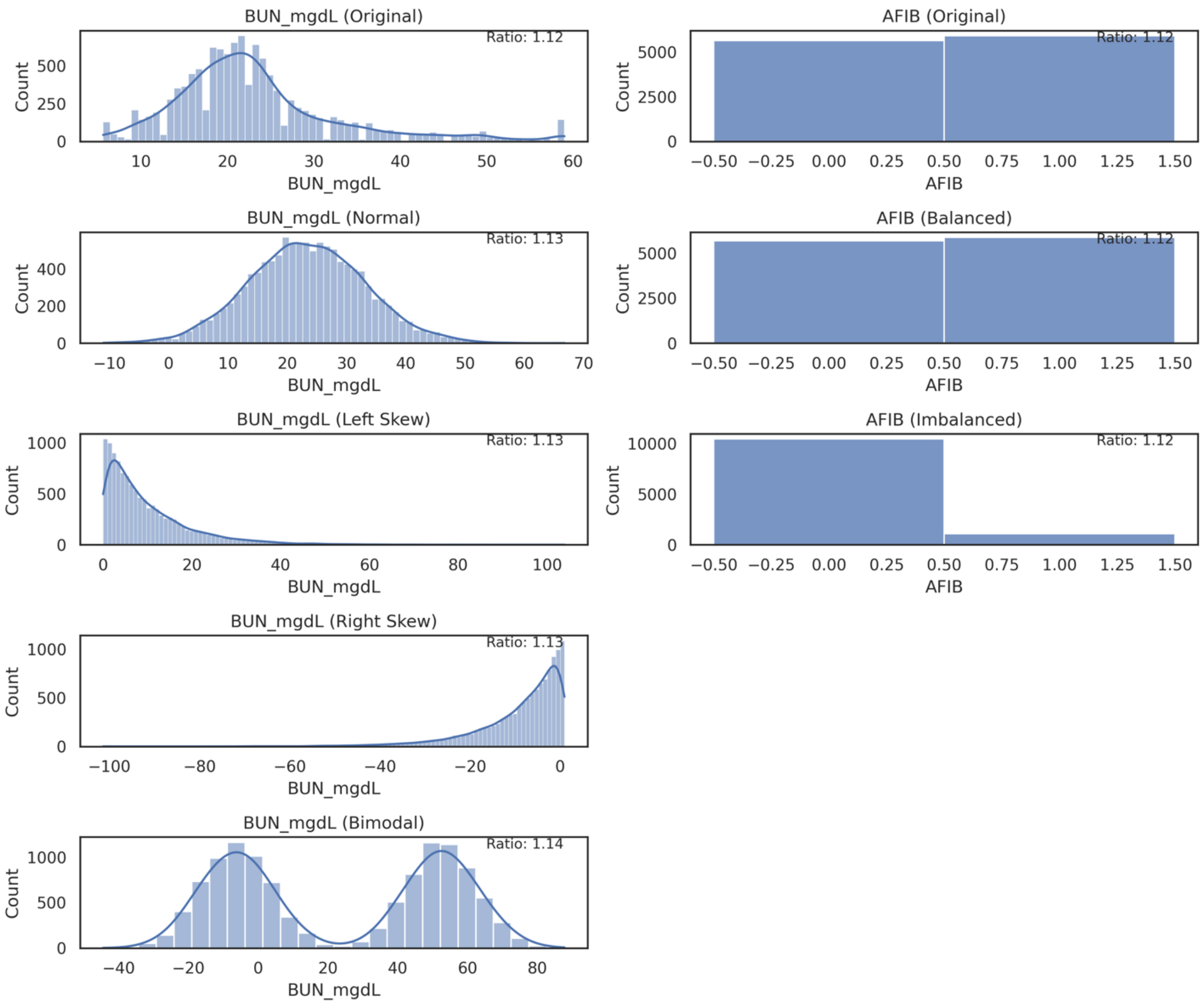

**Figure S13: Simulated Blood Urea Nitrogen Level and Atrial Fibrillation covariate distributions with calculated Phenotypic Distance Metric.** Left column depicts the original Blood Urea Nitrogen level distribution along with normal, skewed, and bimodal transformations with the Phenotypic Distance Metric (ratio) in the upper right hand corner. Right column depicts the original proportion of patients who had (Right bar) or did not have (Left bar) atrial fibrillation, balanced proportion (50/50 split) and an imbalanced (90/10 split) proportion with the Phenotypic Distance Metric (ratio) in the upper right hand corner. Abbreviations: BUN: Blood Urea Nitrogen, AFIB: Atrial Fibrillation.

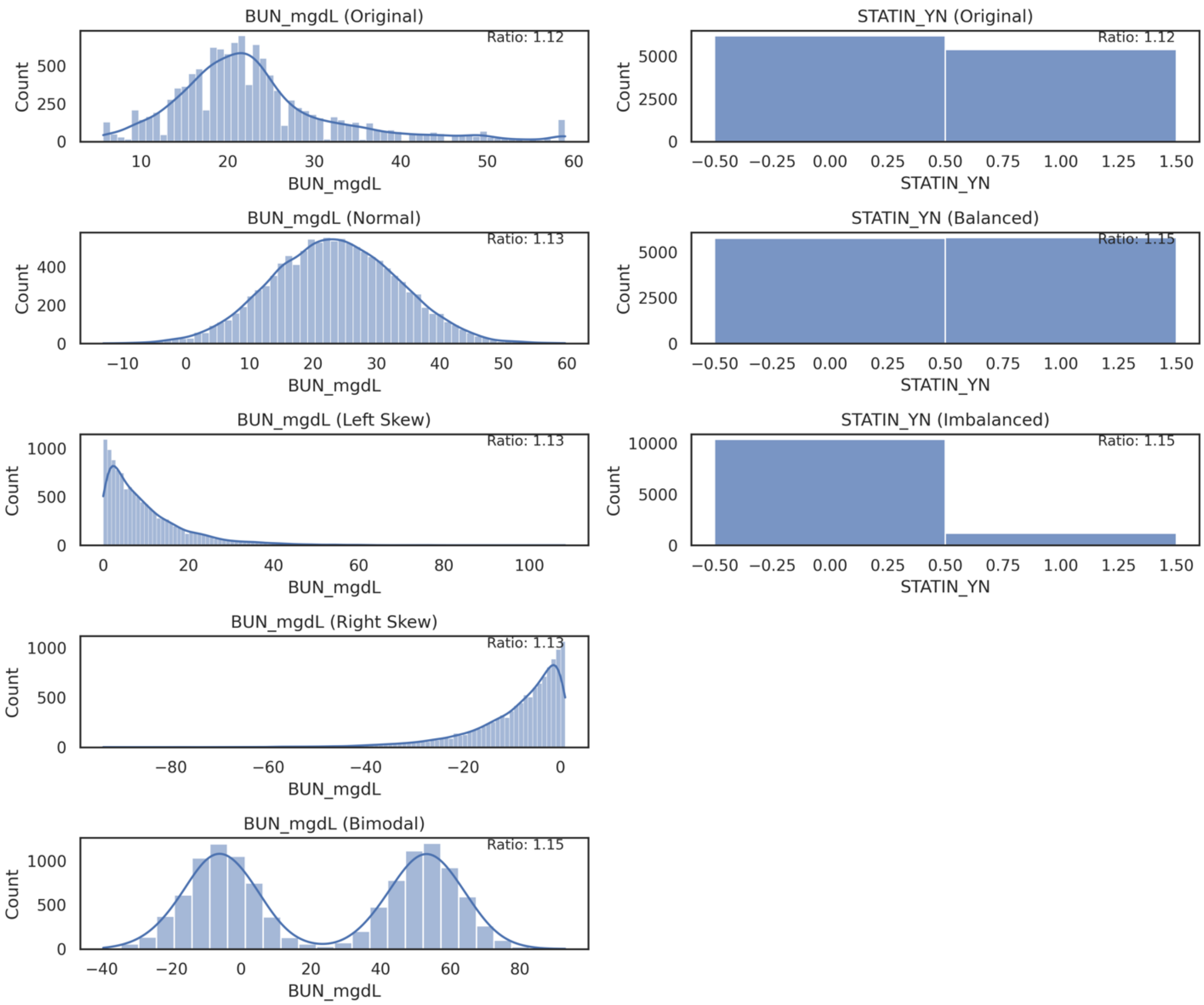

**Figure S14: Simulated Blood Urea Nitrogen Level and Statin Use covariate distributions with calculated Phenotypic Distance Metric.** Left column depicts the original Blood Urea Nitrogen level distribution along with normal, skewed, and bimodal transformations with the Phenotypic Distance Metric (ratio) in the upper right hand corner. Right column depicts the original proportion of patients who used (Right bar) or did not use (Left bar) a statin, balanced proportion (50/50 split) and an imbalanced (90/10 split) proportion with the Phenotypic Distance Metric (ratio) in the upper right hand corner. Abbreviations: BUN: Blood Urea Nitrogen.

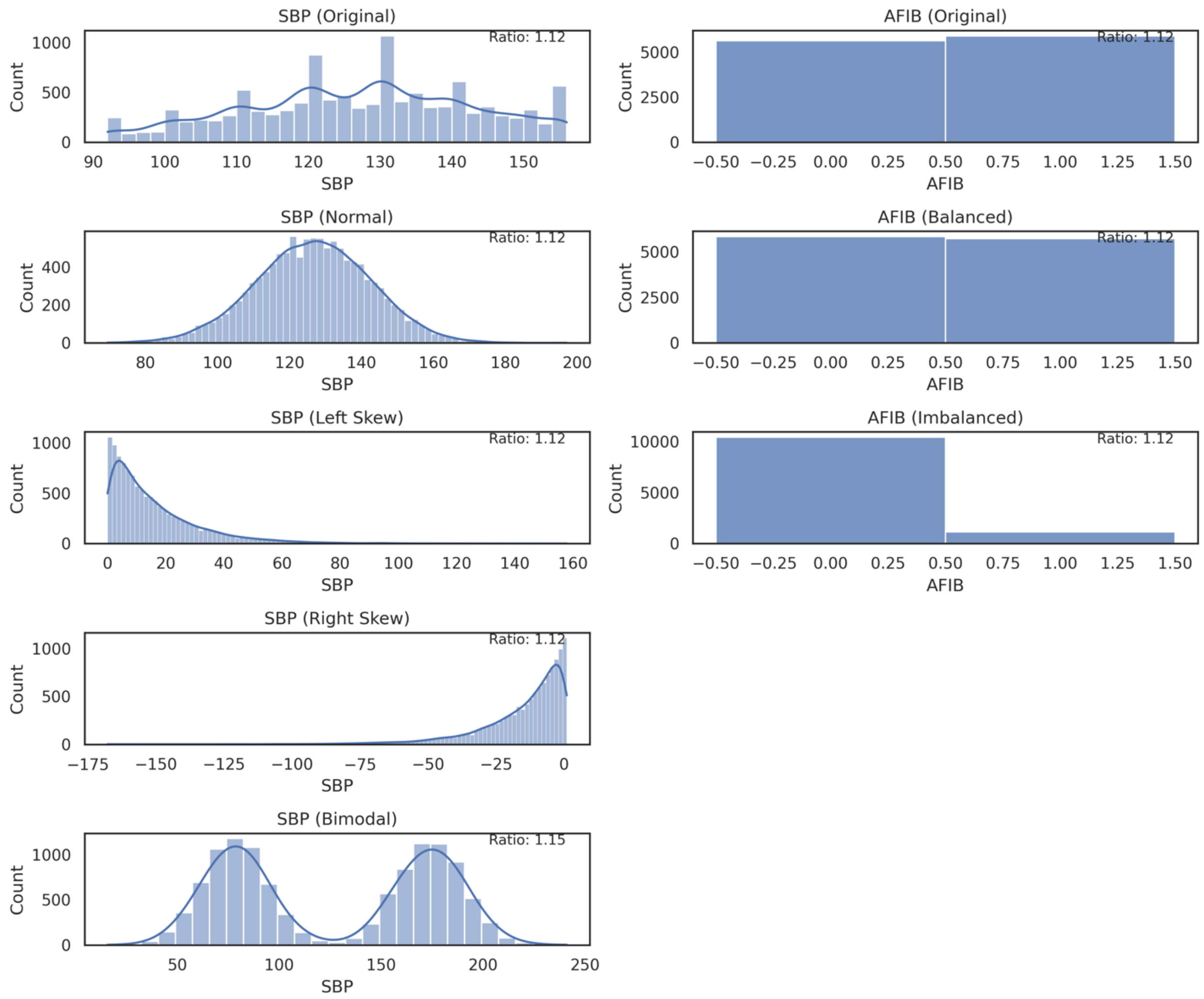

**Figure S15: Simulated Systolic Blood Pressure and Atrial Fibrillation covariate distributions with calculated Phenotypic Distance Metric.** Left column depicts the original Systolic Blood Pressure distribution along with normal, skewed, and bimodal transformations with the Phenotypic Distance Metric (ratio) in the upper right hand corner. Right column depicts the original proportion of patients who had (Right bar) or did not have (Left bar) atrial fibrillation, balanced proportion (50/50 split) and an imbalanced (90/10 split) proportion with the Phenotypic Distance Metric (ratio) in the upper right hand corner. Abbreviations: SBP: Systolic Blood Pressure, AFIB: Atrial Fibrillation.

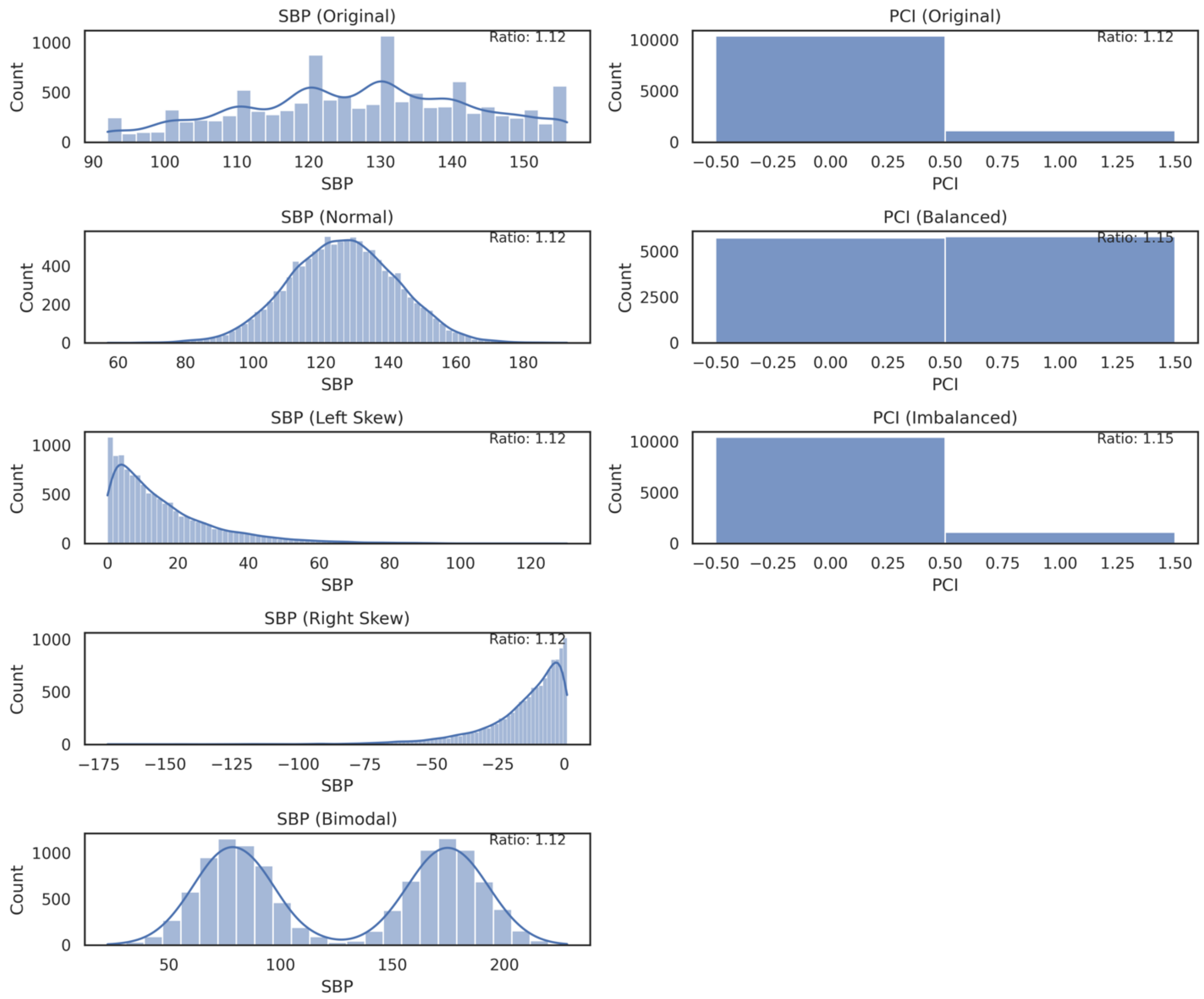

**Figure S16: Simulated Systolic Blood Pressure and Percutaneous Coronary Intervention covariate distributions with calculated Phenotypic Distance Metric.** Left column depicts the original Systolic Blood Pressure distribution along with normal, skewed, and bimodal transformations with the Phenotypic Distance Metric (ratio) in the upper right hand corner. Right column depicts the original proportion of patients who had (Right bar) or did not have (Left bar) atrial fibrillation, balanced proportion (50/50 split) and an imbalanced (90/10 split) proportion with the Phenotypic Distance Metric (ratio) in the upper right hand corner. Abbreviations: SBP: Systolic Blood Pressure, PCI: Percutaneous Coronary Intervention.

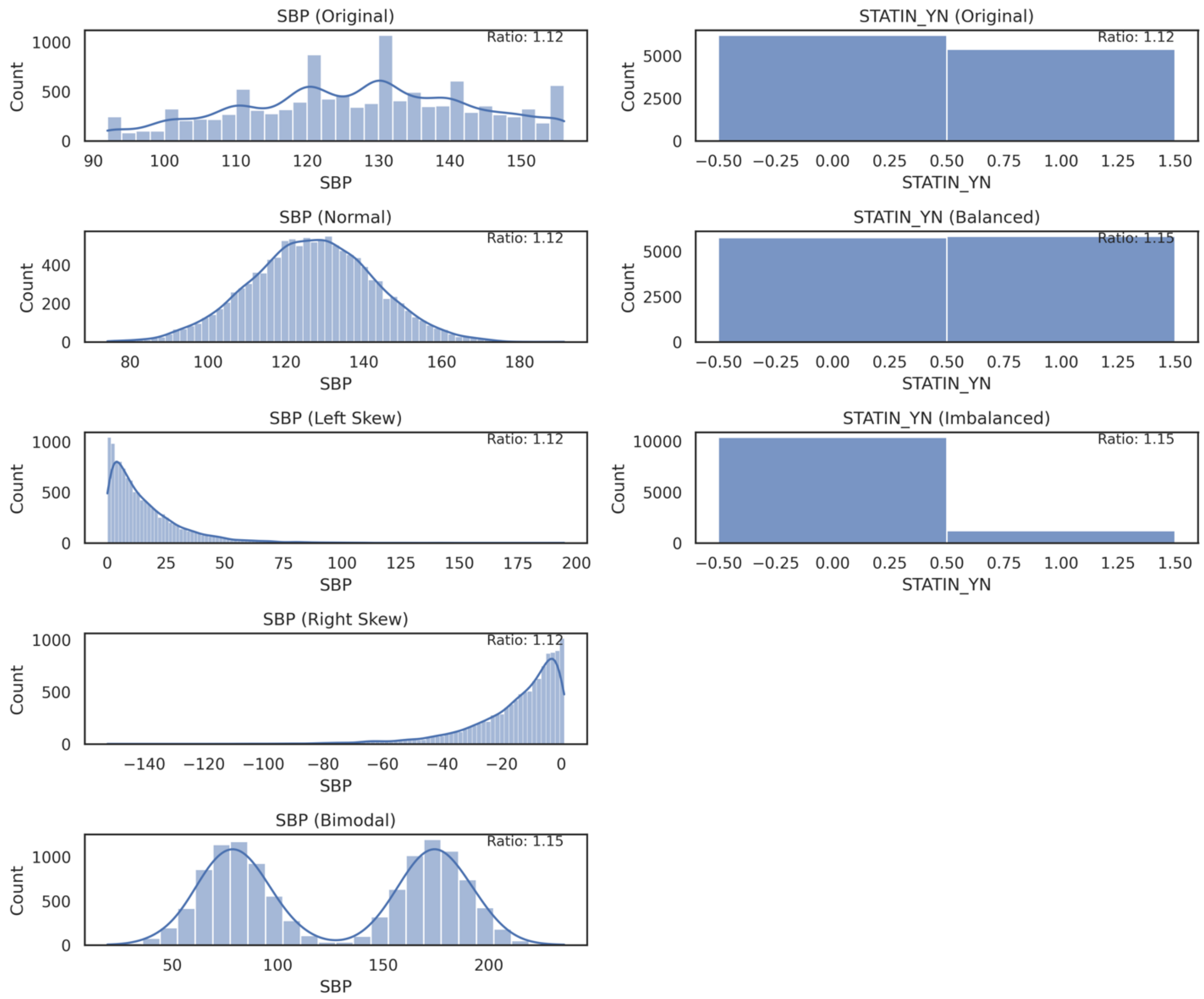

**Figure S17: Simulated Systolic Blood Pressure and Statin Use covariate distributions with calculated Phenotypic Distance Metric.** Left column depicts the original Systolic Blood Pressure distribution along with normal, skewed, and bimodal transformations with the Phenotypic Distance Metric (ratio) in the upper right hand corner. Right column depicts the original proportion of patients who used (Right bar) or did not use (Left bar) a statin, balanced proportion (50/50 split) and an imbalanced (90/10 split) proportion with the Phenotypic Distance Metric (ratio) in the upper right hand corner. Abbreviations: SBP: Systolic Blood Pressure.

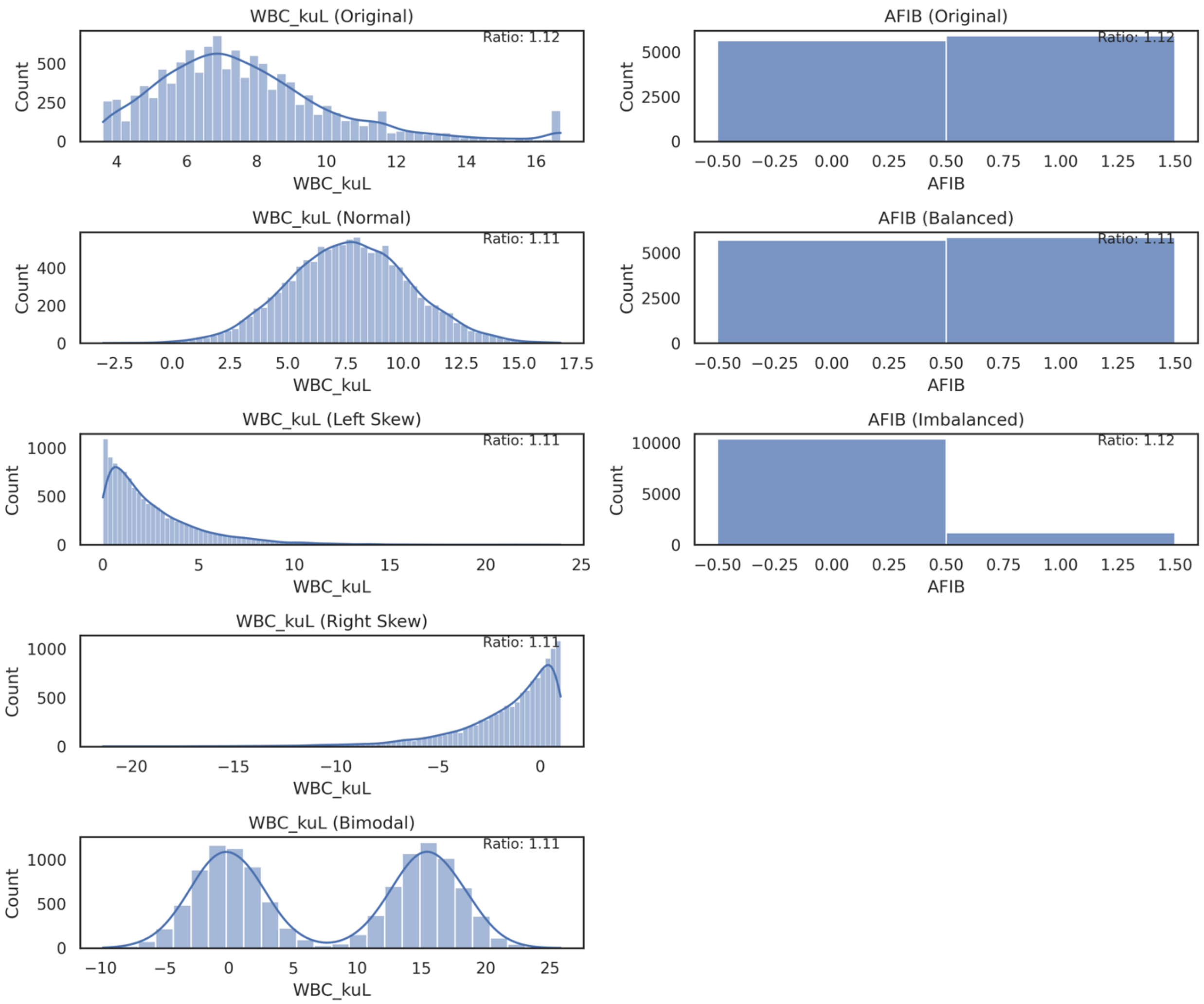

**Figure S18: Simulated White Blood Cell Count and Atrial Fibrillation covariate distributions with calculated Phenotypic Distance Metric.** Left column depicts the original white blood cell count distribution along with normal, skewed, and bimodal transformations with the Phenotypic Distance Metric (ratio) in the upper right hand corner. Right column depicts the original proportion of patients who had (Right bar) or did not have (Left bar) atrial fibrillation, balanced proportion (50/50 split) and an imbalanced (90/10 split) proportion with the Phenotypic Distance Metric (ratio) in the upper right hand corner. Abbreviations: WBC: White Blood Count, AFIB: Atrial Fibrillation.

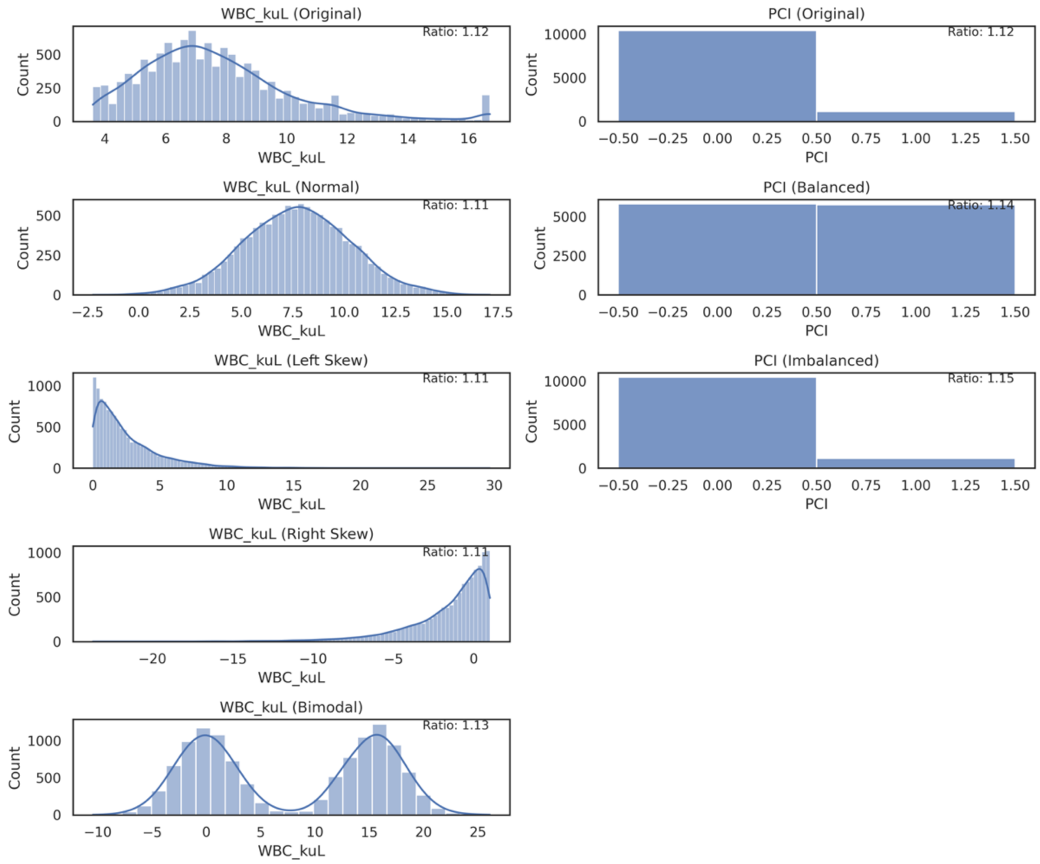

**Figure S19: Simulated White Blood Cell Count and Percutaneous Coronary Intervention covariate distributions with calculated Phenotypic Distance Metric.** Left column depicts the original Systolic Blood Pressure distribution along with normal, skewed, and bimodal transformations with the Phenotypic Distance Metric (ratio) in the upper right hand corner. Right column depicts the original proportion of patients who had (Right bar) or did not have (Left bar) a percutaneous coronary intervention, balanced proportion (50/50 split) and an imbalanced (90/10 split) proportion with the Phenotypic Distance Metric (ratio) in the upper right hand corner. Abbreviations: WBC: White Blood Count, PCI: Percutaneous Coronary Intervention.

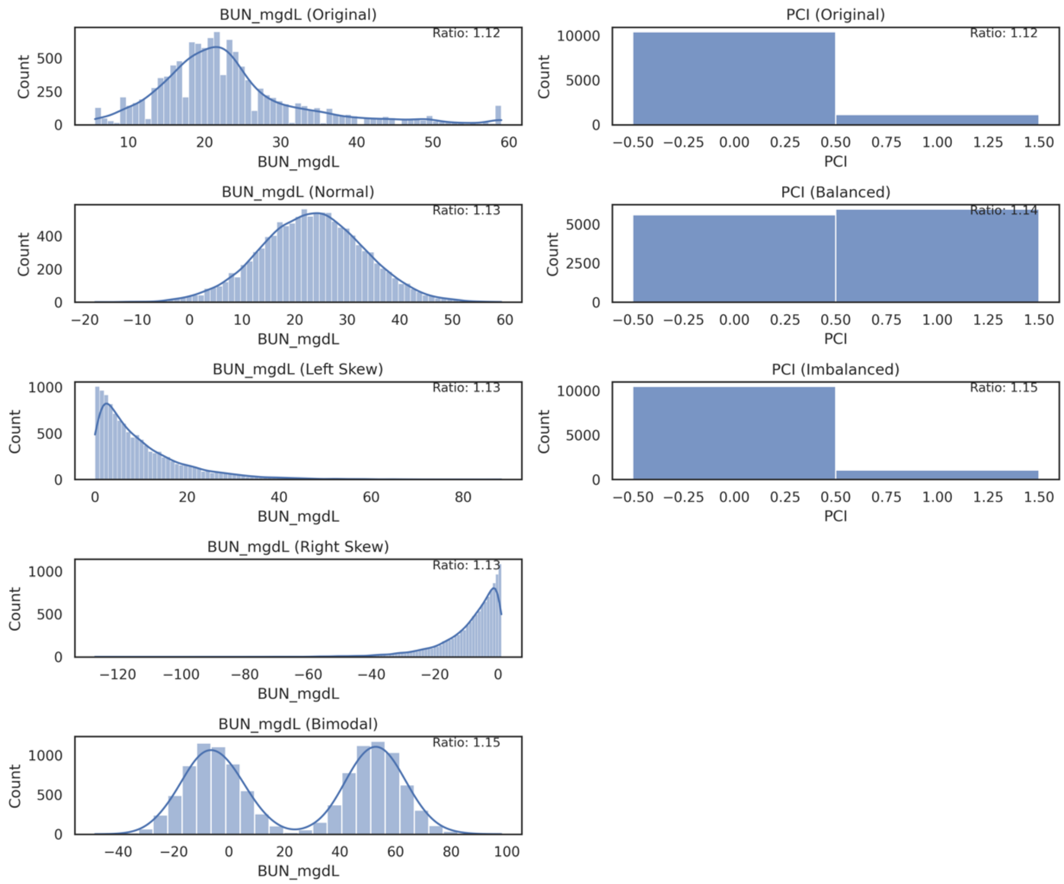

**Figure S20: Simulated Blood Urea Nitrogen Level and Percutaneous Coronary Intervention.** Covariate distributions with calculated Phenotypic Distance Metric. Left column depicts the original Blood Urea Nitrogen level distribution along with normal, skewed, and bimodal transformations with the Phenotypic Distance Metric (ratio) in the upper right hand corner. Right column depicts the original proportion of patients who had (Right bar) or did not have (Left bar) a percutaneous coronary intervention, balanced proportion (50/50 split) and an extreme (90/10 split) proportion with the Phenotypic Distance Metric (ratio) in the upper right hand corner. Abbreviations: BUN: Blood Urea Nitrogen, PCI: Percutaneous Coronary Intervention.

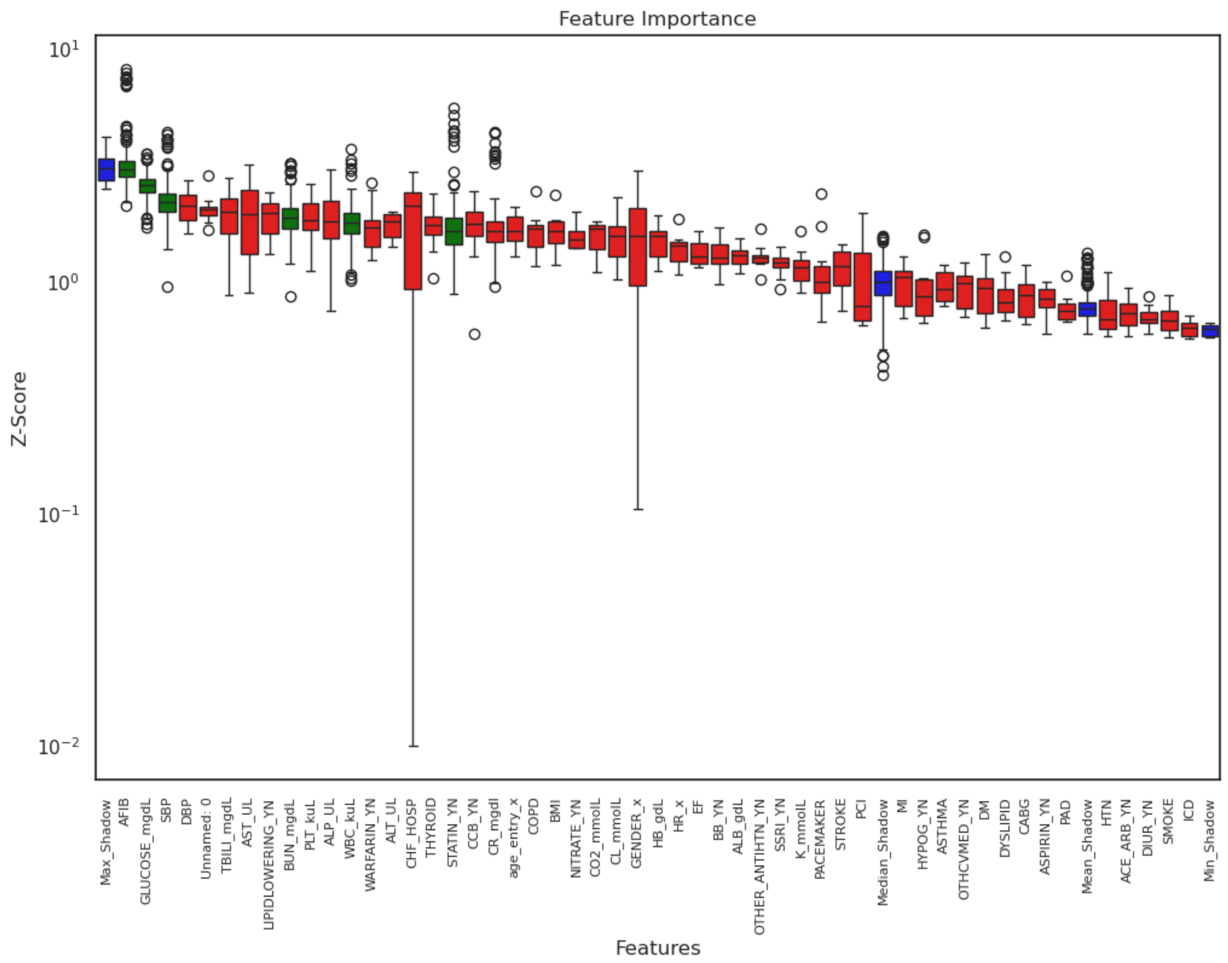

**Figure S21: Features Ranked by importance using XGBoost Model trained on TOPCAT-US participants.** Green boxes are those features deemed to be important for predicting personalized benefit from spironolactone, yellow boxes have uncertain importance, and red boxes have no importance. Blue boxes are synthetic, shuffled features to provide a baseline to compare to real features. Abbreviations: ACE: Angiotensin Converting Enzyme, BH: Bridgeport Hospital,  dL: deciliters, g:grams, GH: Greenwich Hospital, L: Liters, k: thousand, LMH: Lawrence & Memorial Hospital kg: kilograms, $m^3$: meters cubed,  mg: milligrams, mmol: milli-moles, n: number, Q1: 25% percentile, Q3: 75% percentile, SMD: Standardized Mean Difference, SRC: St. Raphael's Campus, U: Units, uL: milliliters, YNHH: Yale New Haven Hospital, YSC: York Street Campus, Alk, Phos: Alkaline Phosphatase, ALT: Alanine Transferase, AST: Aspartate Transferase, BILITOT: Total Bilirubin, BMI: Body Mass Index, BUN: Blood Urea Nitrogen, CL: Chloride, CO2: Bicarbonate, DBP: Diastolic Blood Pressure, EF: Ejection Fraction, GFR: Glomerular Filtration Rate, GLU: Glucose, HCT: Hematocrit, HGB: Hemoglobin, HR: Heart Rate, K: Potassium, NA: Sodium, Plt: Platelets, SBP: Systolic Blood Pressure, WBC: White Blood Cells

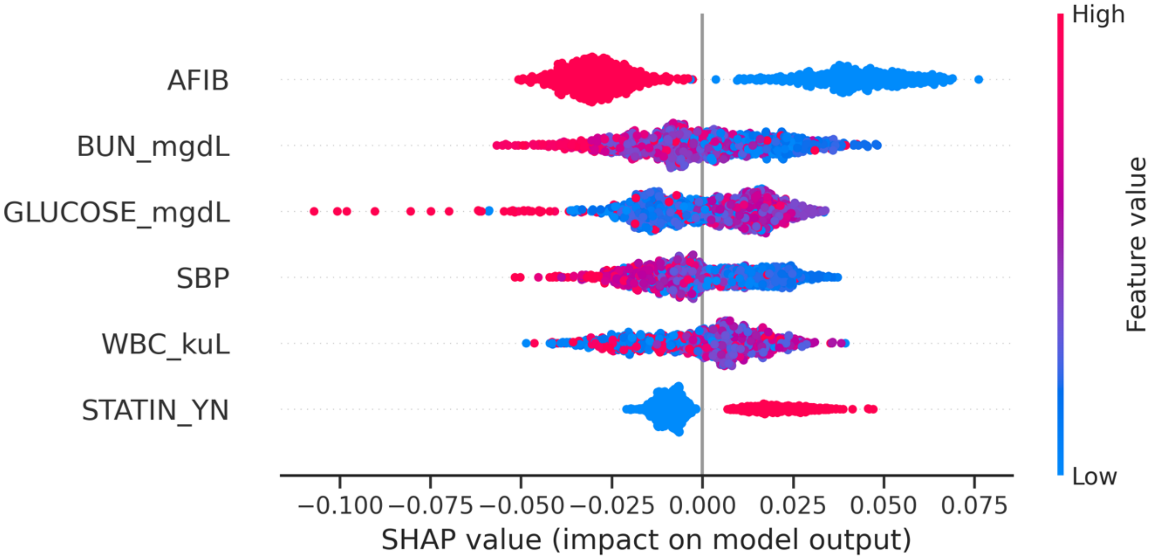

**Figure S22: SHAP analysis train on TOPCAT-US participants showing feature importance for prediction of personalized benefit for spironolactone use.** Y axis are the final features used in the XGBoost model. The x axis quantifies the value of importance of each variable for the model output of that patient. To the left of the grey line means a negative contribution to the model output, and to the right of the grey line means a positive contribution to the model output. Color represents the value of the variable. For binary or categorical variables, red is 1, blue is 0, and for continuous variables, red is a high value while blue is a low value. Abbreviations: AFIB: atrial fibrillation, dL: deciliters, mg: milligrams, uL: milliliters, BUN: Blood Urea Nitrogen, SBP: Systolic Blood Pressure, STATIN_YN: Statin Use, WBC: White Blood Cells

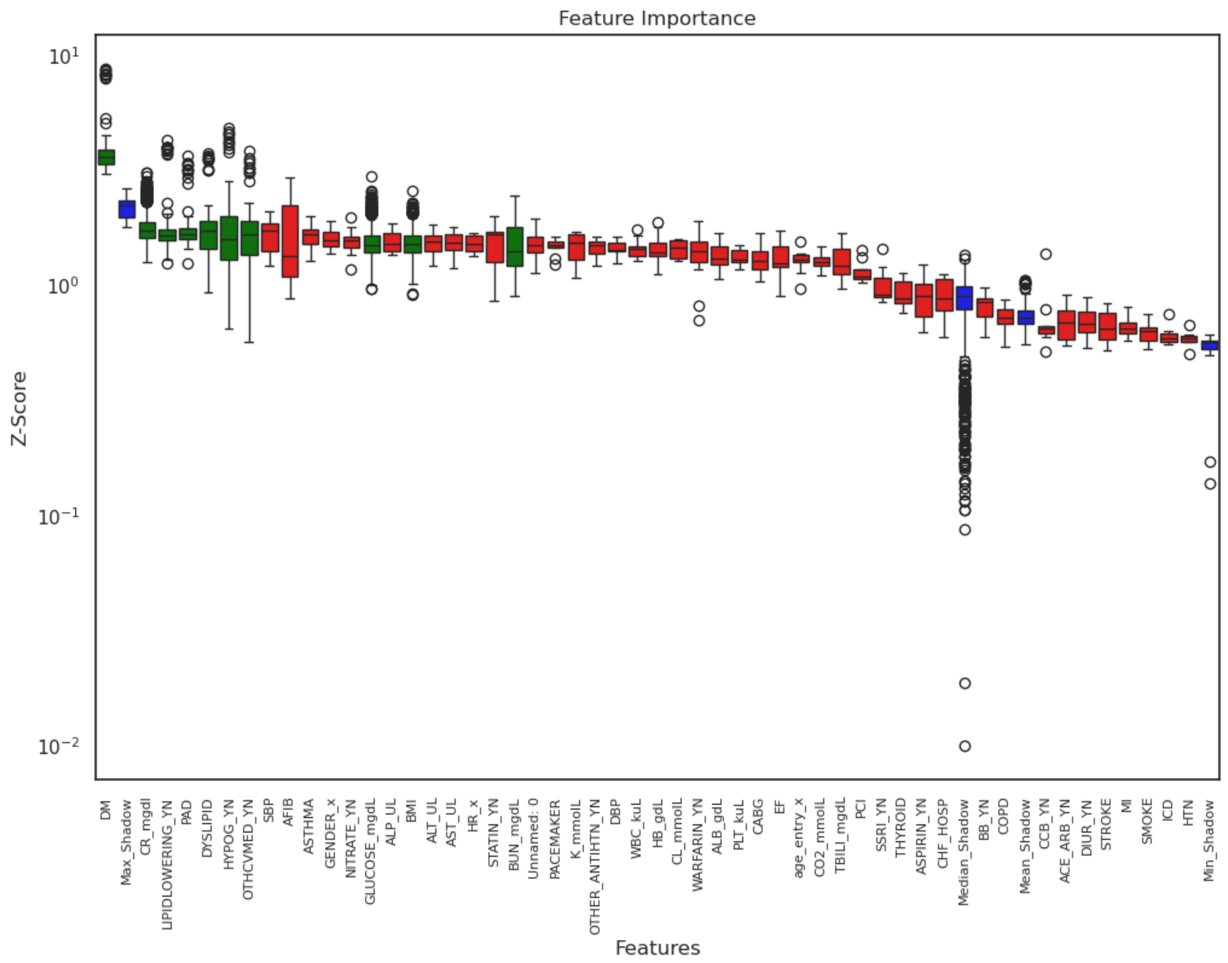

**Figure S23: Features Ranked by importance using XGBoost Model trained on TOPCAT-Eastern-European participants.** Green boxes are those features deemed to be important for predicting personalized benefit from spironolactone, yellow boxes have uncertain importance, and red boxes have no importance. Blue boxes are synthetic, shuffled features to provide a baseline to compare to real features.

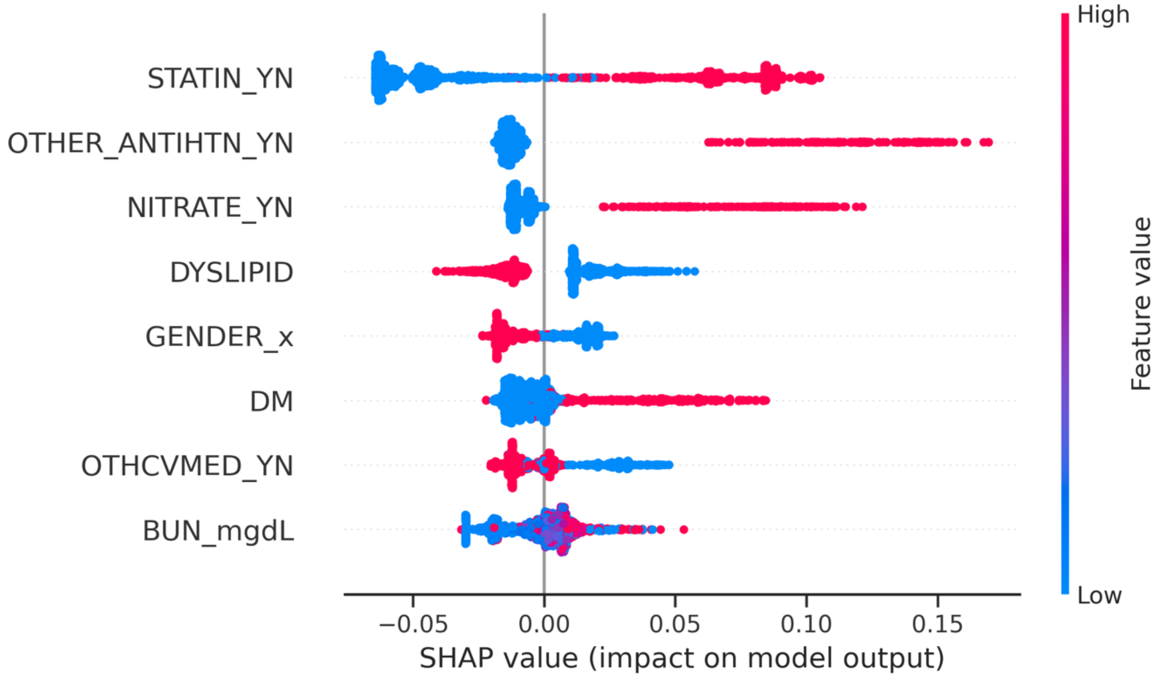

**Figure S24: SHAP analysis train on TOPCAT-Eastern-European participants showing feature importance for prediction of personalized benefit for spironolactone use.** Y axis are the final features used in the XGBoost model. The x axis quantifies the value of importance of each variable for the model output of that patient. To the left of the grey line means a negative contribution to the model output, and to the right of the grey line means a positive contribution to the model output. Color represents the value of the variable. For binary or categorical variables, red is 1, blue is 0, and for continuous variables, red is a high value while blue is a low value. Abbreviations: AFIB: atrial fibrillation, dL: deciliters, mg: milligrams, uL: milliliters, BUN: Blood Urea Nitrogen, SBP: Systolic Blood Pressure, STATIN_YN: Statin Use, WBC: White Blood Cells

| **Redundant Variables** | **Non-covariates** | **Untranslatable to EHR** |
|---|---|---|
| Age of diabetes Mellitus Onset | Documentation Sufficient | Symptom Frequency Score |
| Duration since Diabetes Mellitus diagnosis | How many unopened bottles [of study drug] were returned | Quality of Life Score |
| Paroxysmal atrial fibrillation | Number of bottles dispensed | Symptom Stability Score |
| Chronic atrial fibrillation | Has the subject signed a consent form | Carotid-Femoral Pulse Wave velocity |
| Smoke years | Has the subject consented to blood specimen analysis | Doing yard work |
| Smoke ever | Has the subject consented to urine specimen analysis | How much has heart failure limited life |
| Weight | Time between randomization and visit date | Kansas City Cardiomyopathy Questionnaire |
| Height | Has BNP value | Alcohol use |
| SBP Method | Had hyperkalemia | NYHA class |
| | Has Liver disease | Cardiovascular Exam |
| | On Contraception | Echocardiogram variables |

**Table S1:** Examples of TOPCAT trial variables removed from the study analysis organized by category.

| Diagnosis or Procedure | Column type | Text Search (Not case dependent) |
|---|---|---|
| Atrial Fibrillation | ICD10 List | Starts with “I48.” |
| Asthma | ICD10 List | “Starts with “J45” or “J82.83” |
| Coronary Artery Bypass Graft | ICD10 List | Starts with “Z95.1” |
| Chronic Obstructive Pulmonary Disease | ICD10 List | Starts with “J44” |
| Diabetes Mellitus | ICD10 List | Starts with “E11” |
| Dyslipidemia | ICD10 List | Starts with “E78" |
| Heart Failure | ICD10 List | Starts with “I50” |
| Hypertension | ICD10 List | Starts with “I10” |
| Implantable Cardioverter Defibrillator | Diagnosis Name | contains “ICD” |
| Myocardial Infarction | ICD10 List | Starts with “I21”, “I22”, “I23”, “I24.9”, “I25.9” |
| Pacemaker | Diagnosis Name | “Pacemaker” |
| Peripheral Arterial Disease | ICD10 List | Starts with “I73.9” |
| Percutaneous Coronary Intervention | ICD10 List | contains “percutaneous coronary” |
| Smoking History | ICD10 List | F17 |
| Stroke | ICD10 List | Stroke |
| Thyroid disease | ICD10 List | E0 |
| **Additional Search in Procedure Hospital Encounter** | | |
| Coronary Artery Bypass Graft | Procedure Name | starts with “bypass coronary artery” |
| Implantable Cardioverter Defibrillator | Procedure Name | “icd”, starts with “defib” |
| Pacemaker | Procedure Name | starts with “pace" |
| Percutaneous Coronary Intervention | Procedure Name | starts with “dilation of coronary artery”, “extirpation of matter from coronary artery”, “Introduce Oth Thrombolytic in Coronary Art, Perc”, “Introduce Oth Therap Subst in Coronary Art, Perc”, “Introduce Platelet Inhibitor in Coronary Art, Perc”, “Repair Coronary Artery, One Artery, Percutaneous Endoscopic Approach” |

| Labs | Search type | Search |
|---|---|---|
| Albumin (g/dL) | “Base Name” | “ALBUMIN” |
| Alkaline phosphatase (g/dL) | “Base Name” | “ALKPHOS” |
| Alanine transaminase (IU/L) | “Base Name” | “ALT” |
| Aspartate Transaminase (IU/L) | “Base Name” | “AST” |
| Blood Urea Nitrogen (mg/dL) | “Base Name” | “BUN” |
| Chloride (mmol/L) | “Base Name” | “CL” |
| Bicarbonate (mmol/L) | “Base Name” | “CO2” |
| Creatinine (mg/dL) | “Base Name” | “CREATININE” |
| EGFR | “Base Name” | “EGFR” |
| Glucose (mg/dL) | “Base Name” | “GLU” |
| Potassium (mmol/L) | “Base Name” | “K” |
| Sodium (mmol/L) | “Component Name” | “BKR SODIUM” |
| Total Bilirubin (mg/dL) | “Base Name” | “BILITOT” |
| White Blood Cell Count (k/uL) | “Base Name” | WBC” |
| Hemoglobin (g/dl) | “Base Name” | “HGB” |
| Hematocrit (%) | “Base Name” | “HCT” |
| Platelets (k/uL) | “Base Name” | “PLT” |
| Demographics | | |
| Other | | Not Listed’, ‘Unknown’, ‘Native Hawaiian or Other Pacific Islander’, ‘Middle Eastern or Northern African’, ‘American Indian or Native American’ |
| Asian | | ‘Asian’, ‘Korean’, ‘Asian Indian’, ‘Filipino’ |
| Hispanic or Latino | | ‘Hispanic or Latina/o/x’ or ‘Puerto Rican’ |

**Table S2: Mapping of TOPCAT covariates to EHR definitions.** First column describes the covariate, second column describes the variable used from the EHR tables, and final column describes the text search used. Abbreviations: dL: deciliters, g:grams, L: Liters, k: thousand, kg: kilograms, $m^3$: meters cubed, mg: milligrams, mmol: milli-moles, U: Units, uL: milliliters

| | TOPCAT Variable | EHR variable mapping |
|---|---|---|
| **Text search within "simple generic" medication description** | | |
| | **ACE Inhibitor** | Medication ending with 'pril' |
| | **ARB** | Medication ending with 'sartan' |
| | **Beta Blocker** | 'metoprolol','atenolol','propranolol','pindolol','bisoprolol','acebutolol','lavetalol','carvedilol', or 'bucindolol' |
| | **Calcium Channel Blocker** | Medication ending with 'dipine','pamil', or 'diltiazem' |
| | **Diuretic** | 'chlorthalidone','metolazone','bumetanide','furosemide','torsemide','amiloride','spironolactone','epelernone','triamterene','hydrochlorothiazide','bendroflumethiazide','acetazolamide', or 'dorzolamide' |
| | **Hypoglycemic** | 'tformin','glitazone','glipizide','glimepiride','glyburide','repaglinide','nateglinide','glutide','exenatide','gliptin','gliflozin','acarbose','miglitol', or 'voglibose' |
| | **ACE Inhibitor or ARB** | Medication ending with 'pril' or 'sartan' |
| | **Aspirin** | 'Aspirin' |
| | **Nitrates** | medication starting with 'nitrog','nitrop','isosorbide mononitrate', or 'isosorbide dinitrate' |
| | **Statins** | 'pravastatin', simvastatin','rosuvastatin','lovastatin','atorvastatin','pitavastatin', or 'fluvastatin' |
| | **Warfarin** | 'Warfarin' |
| | **Lipid lowering medications** | 'fibrate','niacin','ezetimibe','lomitapide','lecithin', or 'cholestyramine' |
| | **Other anti-hypertensives** | 'doxazosin','prazosin','terazosin', or 'hydralazine' |
| | **SSRI** | 'Citalopram','Escitalopram','Fluoxetine','Fluvoxamine','Proxetine','Sertraline', or 'Dapoxetine' |
| **Combinations of TOPCAT medication variables and text search within "simple generic" medication description** | | |
| | **Cardiovascular** | Aspirin, Nitrates, Warfarin, or Statins |
| | **Other cardiovascular** | ACE-Is, ARBs, beta blockers, calcium channel blockers, diuretics, lipid lowering medications, other anti-hypertensives,'digoxin','flecainide','procainamide','quinidine','mexiletine','lidocaine','propafenone','sotalol','amiodarone','adenosine','vericiguat' |
| | **Cardiac** | ACE-Is, ARBs, beta blockers, diuretic, cardiovascular, other anti-hypertensives |
| | **Anti-HTN meds** | Beta blockers, ACE-Is, ARBs, Diuretics, other anti-hypertensives, also text search of "simple generic" medications 'urapidil','tolazoline','phenoxygenzamine', or 'phentolamin' |

**Table S3: Mapping of TOPCAT covariate medication definitions to EHR patient datasets.** The top half of the table describes for each medication class (second column) the text searches used in the "Simple Generic" variable of the medication table. The bottom half of the table describes the combinations of the above medication classes used for the covariates described in the second column. Abbreviations: ACE: Angiotensin Converting Enzyme, ARB: Angiontensin Receptor Blocker, HTN: Hypertension, I: Inhibitors

| Covariate | Yes/ No | TOPCAT-US | TOPCAT-Non-US | YNHH SRC | YNHH YSC | BH | GH | LMH | P-Value |
|---|---|---|---|---|---|---|---|---|---|
| **Number** | | 1151 | 2294 | **2482** | **2526** | 1654 | 403 | 1056 | |
| **Demographics** | | | | | | | | | |
| **Female Sex, n (%)** | **No** | 594 (51.6) | 1076 (46.9) | 929 (37.4) | 1159 (45.9) | 699 (42.3) | 176 (43.7) | 467 (44.2) | <0.001 |
| | **Yes** | 557 (48.4) | 1218 (53.1) | 1553 (62.6) | 1367 (54.1) | 955 (57.7) | 227 (56.3) | 589 (55.8) | |
| **Hispanic Ethnicity, n (%)** | **No** | 1096 (95.2) | 2028 (88.4) | 2335 (94.1) | 2383 (94.3) | 1477 (89.3) | 382 (94.8) | 999 (94.6) | <0.001 |
| | **Yes** | 55 (4.8) | 266 (11.6) | 147 (5.9) | 143 (5.7) | 177 (10.7) | 21 (5.2) | 57 (5.4) | |
| **White Race, n (%)** | **No** | 311 (27.0) | 72 (3.1) | 523 (21.1) | 449 (17.8) | 407 (24.6) | 52 (12.9) | 138 (13.1) | <0.001 |
| | **Yes** | 840 (73.0) | 2222 (96.9) | 1959 (78.9) | 2077 (82.2) | 1247 (75.4) | 351 (87.1) | 918 (86.9) | |
| **Black Race, n (%)** | **No** | 885 (76.9) | 2258 (98.4) | 2089 (84.2) | 2232 (88.4) | 1422 (86.0) | 391 (97.0) | 998 (94.5) | <0.001 |
| | **Yes** | 266 (23.1) | 36 (1.6) | 393 (15.8) | 294 (11.6) | 232 (14.0) | 12 (3.0) | 58 (5.5) | |
| **Asian Race, n (%)** | **No** | 1146 (99.6) | 2280 (99.4) | 2463 (99.2) | 2508 (99.3) | 1632 (98.7) | 397 (98.5) | 1048 (99.2) | 0.021 |
| | **Yes** | 5 (0.4) | 14 (0.6) | 19 (0.8) | 18 (0.7) | 22 (1.3) | 6 (1.5) | 8 (0.8) | |
| **Other Race, n (%)** | **No** | 1116 (97.0) | 2259 (98.5) | 2356 (94.9) | 2380 (94.2) | 1488 (90.0) | 367 (91.1) | 981 (92.9) | <0.001 |
| | **Yes** | 35 (3.0) | 35 (1.5) | 126 (5.1) | 146 (5.8) | 166 (10.0) | 36 (8.9) | 75 (7.1) | |
| **Laboratory Values (Units)** | | | | | | | | | |
| **Albumin (g/dL), median [Q1,Q3]** | | 3.9 [3.6,4.2] | 4.2 [3.9,4.5] | 3.8 [3.5,4.0] | 3.8 [3.5,4.1] | 3.8 [3.4,4.1] | 3.4 [3.0,3.7] | 3.4 [3.0,3.7] | <0.001 |
| **Alkaline phosphatase (U/L), median [Q1,Q3]** | | 82.0 [65.0,104.0] | 96.0 [70.0,143.0] | 89.8 [80.3,99.2] | 90.9 [79.0,104.0] | 88.0 [72.0,105.0] | 87.0 [72.0,108.0] | 89.4 [75.0,104.0] | 0.003 |
| **Alanine transaminase (U/L), median [Q1,Q3]** | | 22.0 [16.0,30.5] | 22.0 [16.0,31.0] | 18.0 [14.0,21.4] | 19.0 [15.0,23.1] | 20.0 [15.0,29.0] | 22.0 [17.0,30.0] | 20.8 [17.3,27.0] | <0.001 |
| **Aspartate aminotransferase (U/L), median [Q1,Q3]** | | 23.0 [19.0,30.0] | 23.0 [18.0,28.8] | 22.0 [19.5,25.0] | 22.6 [19.9,27.6] | 23.3 [20.0,30.0] | 22.0 [17.0,28.0] | 21.0 [16.0,24.6] | <0.001 |
| **Bicarbonate (mmol/L), median [Q1,Q3]** | | 28.0 [26.0,30.0] | 28.0 [27.0,28.7] | 26.9 [25.4,27.8] | 26.8 [25.0,28.0] | 27.0 [25.0,29.0] | 28.0 [26.0,31.0] | 28.0 [26.6,30.0] | <0.001 |
| **Blood Urea Nitrogen (mg/dL), median [Q1,Q3]** | | 22.0 [17.0,29.0] | 18.4 [15.4,22.4] | 22.3 [19.0,26.2] | 22.4 [18.0,27.0] | 22.0 [17.0,29.0] | 23.0 [18.0,31.0] | 22.1 [18.0,27.2] | <0.001 |

| | | | | | | | | | |
|---|---|---|---|---|---|---|---|---|---|
| **Chloride (mmol/L), median [Q1,Q3]** | | 102.0 [100.0,105.0] | 103.0 [100.0,106.0] | 102.0 [100.0,103.0] | 102.0 [100.0,103.0] | 102.0 [99.0,104.0] | 104.0 [101.0,107.0] | 102.9 [101.0,106.0] | <0.001 |
| **Creatinine (mg/dL), median [Q1,Q3]** | | 1.1 [0.9,1.4] | 1.0 [0.9,1.2] | 1.1 [0.9,1.3] | 1.1 [0.9,1.3] | 1.0 [0.8,1.3] | 1.1 [0.8,1.4] | 1.1 [0.9,1.4] | <0.001 |
| **Fasting glucose (mg/dL), median [Q1,Q3]** | | 105.0 [91.0,136.0] | 101.8 [90.9,116.4] | 113.0 [102.0,139.0] | 115.0 [102.0,142.0] | 112.8 [99.0,140.6] | 105.0 [93.0,124.5] | 111.0 [98.0,139.9] | <0.001 |
| **Hemoglobin (g/dL), median [Q1,Q3]** | | 12.7 [11.6,13.9] | 13.5 [12.5,14.7] | 11.6 [10.2,13.0] | 11.7 [10.0,12.9] | 11.7 [10.1,13.0] | 11.9 [10.5,13.3] | 11.7 [10.2,13.0] | <0.001 |
| **Platelets (k/uL), median [Q1,Q3]** | | 219.0 [183.0,262.0] | 225.0 [194.0,263.0] | 222.0 [174.0,272.8] | 214.0 [166.0,268.7] | 216.0 [171.1,262.7] | 220.0 [175.0,271.5] | 224.2 [176.0,272.0] | <0.001 |
| **Potassium (mmol/L), median [Q1,Q3]** | | 4.2 [3.9,4.5] | 4.4 [4.0,4.6] | 4.2 [4.1,4.3] | 4.2 [4.0,4.4] | 4.2 [3.9,4.4] | 4.2 [4.0,4.5] | 4.2 [3.9,4.3] | <0.001 |
| **Total Bilirubin (mg/dL), median [Q1,Q3]** | | 0.6 [0.4,0.8] | 0.7 [0.5,0.9] | 0.5 [0.4,0.6] | 0.5 [0.4,0.7] | 0.5 [0.4,0.7] | 0.6 [0.5,0.8] | 0.5 [0.4,0.7] | <0.001 |
| **White Blood Cells (k/uL), median [Q1,Q3]** | | 7.1 [5.9,8.4] | 6.6 [5.5,7.9] | 7.6 [6.2,9.3] | 7.6 [6.1,9.4] | 7.5 [6.0,9.1] | 7.3 [5.8,9.1] | 7.9 [6.4,9.5] | <0.001 |
| **Vital Signs** | | | | | | | | | |
| **Body Mass Index (kg/m$^2$), median [Q1,Q3]** | | 33.5 [28.6,39.5] | 29.9 [26.8,33.9] | 28.8 [24.1,35.5] | 28.5 [24.4,34.6] | 29.1 [24.8,34.6] | 26.3 [22.9,31.0] | 29.2 [24.8,35.2] | <0.001 |
| **Heart Rate (beats/min), median [Q1,Q3]** | | 68.0 [60.0,76.0] | 68.0 [62.0,76.0] | 75.0 [66.0,85.0] | 75.0 [66.0,85.0] | 74.0 [65.0,84.0] | 72.0 [63.0,80.0] | 76.0 [66.0,85.0] | <0.001 |
| **Systolic Blood Pressure (mmHg), median [Q1,Q3]** | | 128.0 [118.0,138.0] | 130.0 [120.0,140.0] | 124.0 [112.0,137.0] | 124.0 [110.0,138.0] | 129.0 [117.0,142.0] | 124.0 [110.0,138.0] | 127.0 [115.0,139.0] | <0.001 |
| **Diastolic Blood Pressure (mmHg), median [Q1,Q3]** | | 70.0 [62.0,79.0] | 80.0 [70.2,85.0] | 70.0 [62.0,77.0] | 70.0 [62.0,78.0] | 69.0 [62.0,77.0] | 68.0 [62.0,76.0] | 69.0 [61.0,77.0] | <0.001 |
| **Ejection Fraction (%), median [Q1,Q3]** | | 57.0 [53.0,61.0] | 56.0 [51.0,61.8] | 62.0 [56.0,67.0] | 62.0 [56.0,67.0] | 61.0 [56.8,65.3] | 59.0 [55.2,63.7] | 63.0 [57.5,68.0] | <0.001 |

| | | | | | | | | | |
|---|---|---|---|---|---|---|---|---|---|
| **Age at hospitalization (years) , median [Q1,Q3]** | | 71.0 [63.0,80.0] | 68.0 [60.0,74.0] | 81.0 [71.0,89.0] | 77.0 [68.0,86.0] | 80.0 [70.0,88.0] | 85.0 [77.0,91.0] | 80.0 [71.0,88.0] | <0.001 |
| **Conditions and Procedures** | | | | | | | | | |
| **Myocardial Infarction, n (%)** | **No** | 915 (79.5) | 1637 (71.4) | 2050 (82.6) | 1972 (78.1) | 1334 (80.7) | 340 (84.4) | 900 (85.2) | <0.001 |
| | **Yes** | 236 (20.5) | 657 (28.6) | 432 (17.4) | 554 (21.9) | 320 (19.3) | 63 (15.6) | 156 (14.8) | |
| **Stroke, n (%)** | **No** | 1046 (90.9) | 2134 (93.0) | 2070 (83.4) | 2135 (84.5) | 1386 (83.8) | 344 (85.4) | 894 (84.7) | <0.001 |
| | **Yes** | 105 (9.1) | 160 (7.0) | 412 (16.6) | 391 (15.5) | 268 (16.2) | 59 (14.6) | 162 (15.3) | |
| **Chronic Obstructive Pulmonary Disease, n (%)** | **No** | 946 (82.2) | 2096 (91.4) | 1550 (62.4) | 1705 (67.5) | 1068 (64.6) | 300 (74.4) | 575 (54.5) | <0.001 |
| | **Yes** | 205 (17.8) | 198 (8.6) | 932 (37.6) | 821 (32.5) | 586 (35.4) | 103 (25.6) | 481 (45.5) | |
| **Asthma, n (%)** | **No** | 1001 (87.0) | 2221 (96.8) | 1971 (79.4) | 2048 (81.1) | 1250 (75.6) | 355 (88.1) | 873 (82.7) | <0.001 |
| | **Yes** | 150 (13.0) | 73 (3.2) | 511 (20.6) | 478 (18.9) | 404 (24.4) | 48 (11.9) | 183 (17.3) | |
| **Hypertension, n (%)** | **No** | 110 (9.6) | 185 (8.1) | 244 (9.8) | 331 (13.1) | 123 (7.4) | 57 (14.1) | 106 (10.0) | <0.001 |
| | **Yes** | 1041 (90.4) | 2109 (91.9) | 2238 (90.2) | 2195 (86.9) | 1531 (92.6) | 346 (85.9) | 950 (90.0) | |
| **Peripheral Arterial Disease, n (%)** | **No** | 1003 (87.1) | 2123 (92.5) | 2189 (88.2) | 2237 (88.6) | 1449 (87.6) | 353 (87.6) | 898 (85.0) | <0.001 |
| | **Yes** | 148 (12.9) | 171 (7.5) | 293 (11.8) | 289 (11.4) | 205 (12.4) | 50 (12.4) | 158 (15.0) | |
| **Dyslipidemia, n (%)** | **No** | 276 (24.0) | 1093 (47.6) | 740 (29.8) | 746 (29.5) | 355 (21.5) | 115 (28.5) | 286 (27.1) | <0.001 |
| | **Yes** | 875 (76.0) | 1201 (52.4) | 1742 (70.2) | 1780 (70.5) | 1299 (78.5) | 288 (71.5) | 770 (72.9) | |
| **Atrial Fibrillation, n (%)** | **No** | 678 (58.9) | 1553 (67.7) | 1098 (44.2) | 1090 (43.2) | 703 (42.5) | 127 (31.5) | 398 (37.7) | <0.001 |
| | **Yes** | 473 (41.1) | 741 (32.3) | 1384 (55.8) | 1436 (56.8) | 951 (57.5) | 276 (68.5) | 658 (62.3) | |
| **Thyroid disease, n (%)** | **No** | 932 (81.0) | 1973 (86.0) | 1647 (66.4) | 1773 (70.2) | 1095 (66.2) | 269 (66.7) | 772 (73.1) | <0.001 |
| | **Yes** | 219 (19.0) | 321 (14.0) | 835 (33.6) | 753 (29.8) | 559 (33.8) | 134 (33.3) | 284 (26.9) | |
| **Diabetes Mellitus, n (%)** | **No** | 603 (52.4) | 1723 (75.1) | 1399 (56.4) | 1432 (56.7) | 892 (53.9) | 284 (70.5) | 620 (58.7) | <0.001 |
| | **Yes** | 548 (47.6) | 571 (24.9) | 1083 (43.6) | 1094 (43.3) | 762 (46.1) | 119 (29.5) | 436 (41.3) | |
| **Smoker, n (%)** | **No** | 1070 (93.0) | 2015 (87.8) | 2172 (87.5) | 2213 (87.6) | 1490 (90.1) | 389 (96.5) | 922 (87.3) | <0.001 |
| | **Yes** | 81 (7.0) | 279 (12.2) | 310 (12.5) | 313 (12.4) | 164 (9.9) | 14 (3.5) | 134 (12.7) | |
| **Medications** | | | | | | | | | |
| **Angiotensin Converting Enzyme-Inhibitor Use, n (%)** | **No** | 582 (50.6) | 608 (26.5) | 2138 (86.1) | 2097 (83.0) | 1363 (82.4) | 367 (91.1) | 882 (83.5) | <0.001 |
| | **Yes** | 569 (49.4) | 1686 (73.5) | 344 (13.9) | 429 (17.0) | 291 (17.6) | 36 (8.9) | 174 (16.5) | |
| | **No** | 812 (70.5) | 1935 (84.4) | 2105 (84.8) | 2128 (84.2) | 1276 (77.1) | 337 (83.6) | 904 (85.6) | <0.001 |

| | | | | | | | | | |
|---|---|---|---|---|---|---|---|---|---|
| **Angiotensin Receptor Blocker Use, n (%)** | **Yes** | 339 (29.5) | 359 (15.6) | 377 (15.2) | 398 (15.8) | 378 (22.9) | 66 (16.4) | 152 (14.4) | |
| **Beta Blocker Use, n (%)** | **No** | 210 (18.2) | 556 (24.2) | 1146 (46.2) | 1117 (44.2) | 684 (41.4) | 202 (50.1) | 528 (50.0) | <0.001 |
| | **Yes** | 941 (81.8) | 1738 (75.8) | 1336 (53.8) | 1409 (55.8) | 970 (58.6) | 201 (49.9) | 528 (50.0) | |
| **Calcium Channel Blocker Use, n (%)** | **No** | 718 (62.4) | 1433 (62.5) | 1597 (64.3) | 1641 (65.0) | 931 (56.3) | 265 (65.8) | 688 (65.2) | <0.001 |
| | **Yes** | 433 (37.6) | 861 (37.5) | 885 (35.7) | 885 (35.0) | 723 (43.7) | 138 (34.2) | 368 (34.8) | |
| **Diuretic Use n (%)** | **No** | 124 (10.8) | 501 (21.8) | 477 (19.2) | 533 (21.1) | 358 (21.6) | 116 (28.8) | 229 (21.7) | <0.001 |
| | **Yes** | 1027 (89.2) | 1793 (78.2) | 2005 (80.8) | 1993 (78.9) | 1296 (78.4) | 287 (71.2) | 827 (78.3) | |
| **Glucose Lowering Medications Use, n (%)** | **No** | 642 (55.8) | 1841 (80.3) | 2040 (82.2) | 2065 (81.7) | 1300 (78.6) | 358 (88.8) | 912 (86.4) | <0.001 |
| | **Yes** | 509 (44.2) | 453 (19.7) | 442 (17.8) | 461 (18.3) | 354 (21.4) | 45 (11.2) | 144 (13.6) | |
| **ACE-Inhibitor or Angiotensin Receptor Blocker Use, n (%)** | **No** | 268 (23.3) | 274 (11.9) | 1790 (72.1) | 1758 (69.6) | 1038 (62.8) | 308 (76.4) | 743 (70.4) | <0.001 |
| | **Yes** | 883 (76.7) | 2020 (88.1) | 692 (27.9) | 768 (30.4) | 616 (37.2) | 95 (23.6) | 313 (29.6) | |
| **Aspirin Use, n (%)** | **No** | 442 (38.4) | 750 (32.7) | 1796 (72.4) | 1723 (68.2) | 1191 (72.0) | 324 (80.4) | 780 (73.9) | <0.001 |
| | **Yes** | 709 (61.6) | 1544 (67.3) | 686 (27.6) | 803 (31.8) | 463 (28.0) | 79 (19.6) | 276 (26.1) | |
| **Nitrate Use, n (%)** | **No** | 946 (82.2) | 1985 (86.5) | 2244 (90.4) | 2225 (88.1) | 1461 (88.3) | 361 (89.6) | 921 (87.2) | <0.001 |
| | **Yes** | 205 (17.8) | 309 (13.5) | 238 (9.6) | 301 (11.9) | 193 (11.7) | 42 (10.4) | 135 (12.8) | |
| **Statin Use, n (%)** | **No** | 340 (29.5) | 1298 (56.6) | 1428 (57.5) | 1347 (53.3) | 833 (50.4) | 261 (64.8) | 679 (64.3) | <0.001 |
| | **Yes** | 811 (70.5) | 996 (43.4) | 1054 (42.5) | 1179 (46.7) | 821 (49.6) | 142 (35.2) | 377 (35.7) | |
| **Warfarin Use, n (%)** | **No** | 773 (67.2) | 1885 (82.2) | 2219 (89.4) | 2146 (85.0) | 1523 (92.1) | 352 (87.3) | 969 (91.8) | <0.001 |
| | **Yes** | 378 (32.8) | 409 (17.8) | 263 (10.6) | 380 (15.0) | 131 (7.9) | 51 (12.7) | 87 (8.2) | |
| **Lipid Lowering Medication Use, n (%)** | **No** | 954 (82.9) | 2248 (98.0) | 2399 (96.7) | 2420 (95.8) | 1584 (95.8) | 396 (98.3) | 1032 (97.7) | <0.001 |
| | **Yes** | 197 (17.1) | 46 (2.0) | 83 (3.3) | 106 (4.2) | 70 (4.2) | 7 (1.7) | 24 (2.3) | |
| **Other Anti-hypertensive Use, n (%)** | **No** | 919 (79.8) | 2054 (89.5) | 2215 (89.2) | 2236 (88.5) | 1491 (90.1) | 367 (91.1) | 943 (89.3) | 0.001 |
| | **Yes** | 232 (20.2) | 240 (10.5) | 267 (10.8) | 290 (11.5) | 163 (9.9) | 36 (8.9) | 113 (10.7) | |
| **Selective Serotonin Reuptake Inhibitor Use, n (%)** | **No** | 953 (82.8) | 2236 (97.5) | 2102 (84.7) | 2164 (85.7) | 1401 (84.7) | 366 (90.8) | 935 (88.5) | <0.001 |
| | **Yes** | 198 (17.2) | 58 (2.5) | 380 (15.3) | 362 (14.3) | 253 (15.3) | 37 (9.2) | 121 (11.5) | |

| | | | | | | | | | |
|---|---|---|---|---|---|---|---|---|---|
| **Cardiovascular Medication Use, n (%)** | **No** | 56 (4.9) | 234 (10.2) | 910 (36.7) | 841 (33.3) | 571 (34.5) | 180 (44.7) | 455 (43.1) | <0.001 |
| | **Yes** | 1095 (95.1) | 2060 (89.8) | 1572 (63.3) | 1685 (66.7) | 1083 (65.5) | 223 (55.3) | 601 (56.9) | |
| **Other Cardiovascular Medication Use, n (%)** | **No** | 397 (34.5) | 1510 (65.8) | 182 (7.3) | 186 (7.4) | 133 (8.0) | 50 (12.4) | 99 (9.4) | <0.001 |
| | **Yes** | 754 (65.5) | 784 (34.2) | 2300 (92.7) | 2340 (92.6) | 1521 (92.0) | 353 (87.6) | 957 (90.6) | |
| **Cardiac Medication Use, n (%)** | **No** | 6 (0.5) | 10 (0.4) | 218 (8.8) | 230 (9.1) | 156 (9.4) | 57 (14.1) | 121 (11.5) | <0.001 |
| | **Yes** | 1145 (99.5) | 2284 (99.6) | 2264 (91.2) | 2296 (90.9) | 1498 (90.6) | 346 (85.9) | 935 (88.5) | |
| **Implantable Cardioversion Defibrillator Placement, n (%)** | **No** | 1114 (96.8) | 2287 (99.7) | 2425 (97.7) | 2434 (96.4) | 1627 (98.4) | 391 (97.0) | 1039 (98.4) | <0.001 |
| | **Yes** | 37 (3.2) | 7 (0.3) | 57 (2.3) | 92 (3.6) | 27 (1.6) | 12 (3.0) | 17 (1.6) | |
| **Pacemaker Placement, n (%)** | **No** | 971 (84.4) | 2205 (96.1) | 2120 (85.4) | 2135 (84.5) | 1410 (85.2) | 324 (80.4) | 908 (86.0) | <0.001 |
| | **Yes** | 180 (15.6) | 89 (3.9) | 362 (14.6) | 391 (15.5) | 244 (14.8) | 79 (19.6) | 148 (14.0) | |
| **Coronary Arterial Bypass Graft Placement, n (%)** | **No** | 905 (78.6) | 2097 (91.4) | 2265 (91.3) | 2187 (86.6) | 1494 (90.3) | 378 (93.8) | 942 (89.2) | <0.001 |
| | **Yes** | 246 (21.4) | 197 (8.6) | 217 (8.7) | 339 (13.4) | 160 (9.7) | 25 (6.2) | 114 (10.8) | |
| **Percutaneous Coronary Intervention, n (%)** | **No** | 899 (78.1) | 2046 (89.2) | 2337 (94.2) | 2277 (90.1) | 1513 (91.5) | 380 (94.3) | 978 (92.6) | <0.001 |
| | **Yes** | 252 (21.9) | 248 (10.8) | 145 (5.8) | 249 (9.9) | 141 (8.5) | 23 (5.7) | 78 (7.4) | |
| **Prior Heart Failure Hospitalization, n (%)** | **No** | 493 (42.8) | 460 (20.1) | 1977 (79.7) | 2147 (85.0) | 1311 (79.3) | 324 (80.4) | 771 (73.0) | <0.001 |
| | **Yes** | 658 (57.2) | 1834 (79.9) | 505 (20.3) | 379 (15.0) | 343 (20.7) | 79 (19.6) | 285 (27.0) | |

**Table S4: Comprehensive Baseline Characteristics of TOPCAT participants and EHR patients used in this study analysis.** Abbreviations: ACE: Angiotensin Converting Enzyme, BH: Bridgeport Hospital, dL: deciliters, g:grams, GH: Greenwich Hospital, L: Liters, k: thousand, LMH: Lawrence & Memorial Hospital kg: kilograms, $m^3$: meters cubed, mg: milligrams, mmol: milli-moles, n: number, Q1: 25% percentile, Q3: 75% percentile, , SRC: St. Raphael's Campus, U: Units, uL: milliliters, YNHH: Yale New Haven Hospital, YSC: York Street Campus

| Covariate | BH, GH | BM, LMH | BH, TOPCAT-Non-US | BH, TOPCAT-US | BH, YNHH SRC | BH, YNHH YSC | GH, LMH | GH, TOPCAT-Non-US | GH, TOPCAT-US | GH, YNHH SRC | GH, YNHH YSC | LMH, TOPCAT-Non-US | LMH, TOPCAT-US | LMH, YNHH SRC | LMH, YNHH YSC | TOPCAT-Non-US, TOPCAT-US | TOPCAT-Non-US, YNHH, SRC | TOPCAT-Non-US, YNHH, YSC | TOPCAT -US, YNHH, SRC | TOPCAT-US, YNHH, YSC | YNHH SRC, YNHH YSC |
|---|---|---|---|---|---|---|---|---|---|---|---|---|---|---|---|---|---|---|---|---|---|
| | **Cohort Pair** | | | | | | | | | | | | | | | | | | | | |
| **Demographics** | | | | | | | | | | | | | | | | | | | | | |
| **Female Sex** | 0.029 | 0.04 | 0.094 | 0.188 | 0.099 | 0.073 | 0.011 | 0.065 | 0.159 | 0.127 | 0.044 | 0.054 | 0.148 | 0.139 | 0.033 | 0.094 | 0.193 | 0.02 | 0.288 | 0.115 | 0.172 |
| **Hispanic Ethnicity** | 0.204 | 0.196 | 0.028 | 0.223 | 0.174 | 0.185 | 0.008 | 0.232 | 0.02 | 0.031 | 0.02 | 0.224 | 0.028 | 0.023 | 0.012 | 0.251 | 0.202 | 0.213 | 0.051 | 0.04 | 0.011 |
| **White Race** | 0.303 | 0.298 | 0.653 | 0.055 | 0.084 | 0.168 | 0.005 | 0.365 | 0.359 | 0.219 | 0.136 | 0.37 | 0.354 | 0.214 | 0.131 | 0.708 | 0.572 | 0.493 | 0.14 | 0.223 | 0.083 |
| **Black Race,** | 0.404 | 0.291 | 0.478 | 0.235 | 0.051 | 0.071 | 0.125 | 0.095 | 0.626 | 0.452 | 0.337 | 0.214 | 0.52 | 0.34 | 0.221 | 0.693 | 0.523 | 0.414 | 0.185 | 0.306 | 0.122 |
| **Asian Race** | 0.013 | 0.056 | 0.073 | 0.096 | 0.055 | 0.061 | 0.069 | 0.086 | 0.108 | 0.069 | 0.074 | 0.018 | 0.042 | 0.001 | 0.005 | 0.024 | 0.019 | 0.013 | 0.043 | 0.037 | 0.006 |
| **Other Race** | 0.038 | 0.105 | 0.371 | 0.286 | 0.188 | 0.158 | 0.067 | 0.337 | 0.25 | 0.152 | 0.121 | 0.277 | 0.186 | 0.085 | 0.054 | 0.102 | 0.2 | 0.228 | 0.103 | 0.134 | 0.031 |
| **Laboratory Values** | | | | | | | | | | | | | | | | | | | | | |
| **Albumin** | -0.707 | -0.718 | 1.035 | 0.368 | 0.117 | 0.109 | -0.002 | 1.764 | 1.141 | 0.884 | 0.858 | 1.787 | 1.159 | 0.899 | 0.871 | -0.758 | -1.007 | -0.987 | -0.275 | -0.273 | -0.006 |
| **Alk. Phos.** | -0.005 | 0.009 | 0.386å | -0.178 | -0.017 | 0.076 | 0.015 | 0.394 | -0.175 | -0.012 | 0.083 | 0.388 | -0.195 | -0.028 | 0.07 | -0.521 | -0.42 | -0.333 | 0.179 | 0.258 | 0.102 |
| **ALT** | 0.178 | 0.06 | 0.113 | 0.11 | -0.369 | -0.194 | -0.127 | -0.068 | -0.071 | -0.573 | -0.387 | 0.058 | 0.055 | -0.463 | -0.27 | -0.003 | -0.504 | -0.318 | -0.503 | -0.317 | 0.182 |
| **AST** | -0.205 | -0.384 | -0.183 | -0.102 | -0.291 | -0.094 | -0.177 | 0.028 | 0.11 | -0.068 | 0.112 | 0.212 | 0.295 | 0.125 | 0.291 | 0.085 | -0.102 | 0.087 | -0.192 | 0.006 | 0.19 |
| **Bicarbonate** | 0.343 | 0.4 | 0.284 | 0.401 | -0.111 | -0.123 | 0.045 | -0.136 | 0.038 | -0.5 | -0.504 | -0.201 | -0.009 | -0.574 | -0.576 | 0.199 | -0.489 | -0.491 | -0.583 | -0.585 | -0.015 |

| | | | | | | | | | | | | | | | | | | | | | |
|---|---|---|---|---|---|---|---|---|---|---|---|---|---|---|---|---|---|---|---|---|---|
| **BUN** | 0.109 | -0.007 | -0.57 | -0.041 | -0.023 | -0.014 | -0.12 | -0.691 | -0.153 | -0.138 | -0.127 | -0.589 | -0.035 | -0.016 | -0.007 | 0.545 | 0.59 | 0.581 | 0.02 | 0.028 | 0.009 |
| **Chloride** | 0.502 | 0.417 | 0.344 | 0.257 | 0.023 | 0.003 | -0.109 | -0.16 | -0.289 | -0.52 | -0.525 | -0.058 | -0.186 | -0.431 | -0.438 | -0.117 | -0.35 | -0.359 | -0.259 | -0.271 | -0.021 |
| **Creatinine** | 0.087 | 0.17 | -0.195 | 0.222 | 0.103 | 0.098 | 0.076 | -0.284 | 0.12 | 0.009 | 0.005 | -0.393 | 0.041 | -0.072 | -0.076 | 0.478 | 0.325 | 0.318 | -0.119 | -0.124 | -0.005 |
| **Fasting glucose** | -0.268 | -0.017 | -0.396 | -0.107 | -0.019 | 0.019 | 0.248 | -0.135 | 0.145 | 0.263 | 0.301 | -0.375 | -0.089 | -0.001 | 0.037 | 0.266 | 0.399 | 0.435 | 0.093 | 0.129 | 0.04 |
| **Hemoglobin** | 0.113 | 0.028 | 1.112 | 0.644 | -0.013 | -0.043 | -0.086 | 1.001 | 0.527 | -0.126 | -0.154 | 1.089 | 0.618 | -0.04 | -0.07 | -0.531 | -1.12 | -1.129 | -0.655 | -0.674 | -0.03 |
| **Platelets** | 0.102 | 0.09 | 0.149 | 0.05 | 0.089 | 0.006 | -0.013 | 0.03 | -0.063 | -0.012 | -0.093 | 0.046 | -0.049 | 0.001 | -0.081 | -0.11 | -0.044 | -0.136 | 0.049 | -0.041 | -0.081 |
| **Potassium** | 0.078 | -0.163 | 0.351 | 0.018 | 0.095 | 0.089 | -0.239 | 0.27 | -0.059 | 0.01 | 0.006 | 0.51 | 0.178 | 0.271 | 0.263 | -0.326 | -0.282 | -0.282 | 0.074 | 0.068 | -0.005 |
| **Total Bilirubin** | 0.151 | -0.035 | 0.357 | 0.051 | -0.213 | -0.081 | -0.197 | 0.211 | -0.105 | -0.394 | -0.241 | 0.421 | 0.091 | -0.191 | -0.05 | -0.32 | -0.637 | -0.459 | -0.283 | -0.138 | 0.132 |
| **WBCs** | -0.062 | 0.116 | -0.474 | -0.267 | 0.045 | 0.065 | 0.178 | -0.397 | -0.194 | 0.106 | 0.124 | -0.615 | -0.404 | -0.069 | -0.045 | 0.251 | 0.515 | 0.52 | 0.313 | 0.326 | 0.021 |
| **Vital Signs** | | | | | | | | | | | | | | | | | | | | | |
| **BMI** | -0.364 | 0.027 | 0.043 | 0.525 | 0.013 | -0.042 | 0.384 | 0.473 | 0.92 | 0.36 | 0.315 | 0.011 | 0.486 | -0.014 | -0.068 | 0.559 | -0.027 | -0.091 | -0.49 | -0.561 | -0.053 |
| **Heart Rate** | -0.178 | 0.061 | -0.562 | -0.553 | 0.041 | 0.039 | 0.241 | -0.365 | -0.361 | 0.223 | 0.222 | -0.641 | -0.629 | -0.021 | -0.023 | -0.009 | 0.627 | 0.627 | 0.615 | 0.615 | -0.002 |
| **SBP** | -0.187 | -0.091 | 0.145 | -0.06 | -0.216 | -0.207 | 0.099 | 0.356 | 0.139 | -0.028 | -0.02 | 0.252 | 0.036 | -0.128 | -0.119 | -0.227 | -0.392 | -0.378 | -0.17 | -0.16 | 0.007 |
| **DBP** | -0.065 | -0.034 | 0.943 | 0.091 | 0.012 | 0.021 | 0.028 | 1.038 | 0.156 | 0.078 | 0.086 | 0.944 | 0.121 | 0.046 | 0.054 | -0.824 | -0.936 | -0.911 | -0.079 | -0.069 | 0.009 |

| | | | | | | | | | | | | | | | | | | | | | |
|---|---|---|---|---|---|---|---|---|---|---|---|---|---|---|---|---|---|---|---|---|---|
| **Ejection Fraction** | -0.197 | 0.257 | -0.593 | -0.461 | 0.084 | 0.087 | 0.448 | -0.415 | -0.282 | 0.27 | 0.274 | -0.81 | -0.685 | -0.163 | -0.161 | 0.125 | 0.639 | 0.645 | 0.516 | 0.52 | 0.003 |
| **Age** | 0.404 | 0.005 | -1.095 | -0.713 | 0.053 | -0.198 | -0.411 | -1.662 | -1.217 | -0.343 | -0.618 | -1.138 | -0.741 | 0.05 | -0.209 | 0.391 | 1.142 | 0.875 | 0.763 | 0.5 | -0.25 |
| **Conditions and Procedures** | | | | | | | | | | | | | | | | | | | | | |
| **MI** | 0.098 | 0.122 | 0.219 | 0.029 | 0.05 | 0.064 | 0.024 | 0.317 | 0.127 | 0.048 | 0.162 | 0.341 | 0.151 | 0.072 | 0.186 | 0.19 | 0.269 | 0.155 | 0.079 | 0.035 | 0.114 |
| **Stroke** | 0.043 | 0.024 | 0.291 | 0.214 | 0.011 | 0.02 | 0.02 | 0.249 | 0.171 | 0.054 | 0.023 | 0.268 | 0.191 | 0.034 | 0.004 | 0.079 | 0.302 | 0.272 | 0.225 | 0.194 | 0.031 |
| **COPD** | 0.216 | 0.207 | 0.683 | 0.407 | 0.044 | 0.062 | 0.427 | 0.461 | 0.189 | 0.26 | 0.153 | 0.913 | 0.625 | 0.163 | 0.27 | 0.274 | 0.731 | 0.618 | 0.452 | 0.344 | 0.106 |
| **Asthma** | 0.329 | 0.175 | 0.647 | 0.295 | 0.092 | 0.134 | 0.154 | 0.335 | 0.034 | 0.237 | 0.195 | 0.48 | 0.12 | 0.083 | 0.041 | 0.367 | 0.558 | 0.519 | 0.203 | 0.161 | 0.042 |
| **Hypertension** | 0.217 | 0.092 | 0.023 | 0.076 | 0.085 | 0.188 | 0.126 | 0.194 | 0.142 | 0.133 | 0.03 | 0.069 | 0.016 | 0.007 | 0.096 | 0.053 | 0.062 | 0.164 | 0.009 | 0.112 | 0.103 |
| **PAD** | 0.001 | 0.075 | 0.166 | 0.014 | 0.018 | 0.029 | 0.074 | 0.166 | 0.014 | 0.018 | 0.03 | 0.24 | 0.061 | 0.093 | 0.104 | 0.18 | 0.148 | 0.137 | 0.032 | 0.043 | 0.011 |
| **Dyslipidemia** | 0.164 | 0.131 | 0.573 | 0.06 | 0.192 | 0.186 | 0.032 | 0.401 | 0.104 | 0.028 | 0.022 | 0.435 | 0.071 | 0.061 | 0.054 | 0.509 | 0.372 | 0.379 | 0.132 | 0.126 | 0.006 |
| **Atrial Fibrillation** | 0.229 | 0.098 | 0.524 | 0.333 | 0.035 | 0.013 | 0.13 | 0.776 | 0.572 | 0.265 | 0.242 | 0.63 | 0.434 | 0.133 | 0.111 | 0.183 | 0.486 | 0.51 | 0.297 | 0.319 | 0.022 |
| **Thyroid disease** | 0.012 | 0.151 | 0.477 | 0.34 | 0.003 | 0.086 | 0.139 | 0.465 | 0.328 | 0.008 | 0.074 | 0.324 | 0.188 | 0.147 | 0.065 | 0.136 | 0.474 | 0.39 | 0.336 | 0.253 | 0.082 |
| **Diabetes Mellitus** | 0.346 | 0.097 | 0.454 | 0.031 | 0.049 | 0.056 | 0.248 | 0.104 | 0.378 | 0.296 | 0.289 | 0.354 | 0.128 | 0.047 | 0.041 | 0.486 | 0.403 | 0.396 | 0.08 | 0.086 | 0.007 |
| **Smoker** | 0.26 | 0.088 | 0.072 | 0.103 | 0.082 | 0.079 | 0.343 | 0.328 | 0.16 | 0.337 | 0.335 | 0.016 | 0.19 | 0.006 | 0.009 | 0.175 | 0.01 | 0.007 | 0.184 | 0.182 | 0.003 |
| **Medications** | | | | | | | | | | | | | | | | | | | | | |
| **Angiotensin Converting Enzyme- Inhibitor Use** | 0.257 | 0.03 | 1.356 | 0.717 | 0.103 | 0.016 | 0.228 | 1.738 | 0.995 | 0.156 | 0.241 | 1.399 | 0.749 | 0.073 | 0.014 | 0.51 | 1.505 | 1.379 | 0.828 | 0.734 | 0.087 |
| **Angiotensin Receptor Blocker Use** | 0.164 | 0.219 | 0.183 | 0.151 | 0.196 | 0.181 | 0.055 | 0.02 | 0.315 | 0.033 | 0.017 | 0.035 | 0.37 | 0.022 | 0.038 | 0.335 | 0.013 | 0.003 | 0.348 | 0.332 | 0.016 |
| **Beta Blocker Use** | 0.177 | 0.174 | 0.371 | 0.522 | 0.097 | 0.058 | 0.002 | 0.556 | 0.714 | 0.079 | 0.118 | 0.553 | 0.711 | 0.077 | 0.116 | 0.147 | 0.472 | 0.431 | 0.626 | 0.584 | 0.039 |
| **Calcium Channel Blocker Use** | 0.195 | 0.182 | 0.126 | 0.124 | 0.165 | 0.178 | 0.013 | 0.069 | 0.07 | 0.03 | 0.017 | 0.056 | 0.058 | 0.017 | 0.004 | 0.002 | 0.039 | 0.052 | 0.041 | 0.054 | 0.013 |

| | | | | | | | | | | | | | | | | | | | | | |
|---|---|---|---|---|---|---|---|---|---|---|---|---|---|---|---|---|---|---|---|---|---|
| **Diuretic Use** | 0.165 | 0.001 | 0.005 | 0.298 | 0.06 | 0.013 | 0.164 | 0.16 | 0.464 | 0.225 | 0.178 | 0.004 | 0.299 | 0.061 | 0.014 | 0.303 | 0.065 | 0.018 | 0.238 | 0.285 | 0.047 |
| **Glucose Lowering Medications Use** | 0.28 | 0.205 | 0.041 | 0.501 | 0.091 | 0.079 | 0.075 | 0.239 | 0.795 | 0.19 | 0.201 | 0.164 | 0.717 | 0.115 | 0.126 | 0.544 | 0.05 | 0.038 | 0.596 | 0.584 | 0.011 |
| **ACE-Inhibitor or Angiotensin Receptor Blocker Use** | 0.3 | 0.162 | 1.234 | 0.869 | 0.201 | 0.145 | 0.138 | 1.707 | 1.255 | 0.099 | 0.154 | 1.475 | 1.07 | 0.039 | 0.017 | 0.301 | 1.538 | 1.449 | 1.121 | 1.048 | 0.056 |
| **Aspirin Use** | 0.198 | 0.042 | 0.856 | 0.718 | 0.008 | 0.083 | 0.156 | 1.098 | 0.946 | 0.19 | 0.282 | 0.906 | 0.765 | 0.034 | 0.125 | 0.119 | 0.866 | 0.76 | 0.727 | 0.626 | 0.091 |
| **Nitrate Use** | 0.04 | 0.034 | 0.054 | 0.174 | 0.068 | 0.008 | 0.074 | 0.094 | 0.213 | 0.028 | 0.047 | 0.02 | 0.14 | 0.101 | 0.026 | 0.12 | 0.122 | 0.047 | 0.241 | 0.166 | 0.075 |
| **Statin Use** | 0.295 | 0.285 | 0.125 | 0.435 | 0.144 | 0.059 | 0.01 | 0.168 | 0.754 | 0.149 | 0.234 | 0.158 | 0.743 | 0.139 | 0.224 | 0.568 | 0.019 | 0.065 | 0.589 | 0.498 | 0.085 |
| **Warfarin Use** | 0.156 | 0.012 | 0.299 | 0.651 | 0.092 | 0.225 | 0.145 | 0.144 | 0.496 | 0.064 | 0.069 | 0.288 | 0.639 | 0.081 | 0.213 | 0.35 | 0.208 | 0.075 | 0.56 | 0.426 | 0.133 |
| **Lipid Lowering Medication Use** | 0.147 | 0.111 | 0.128 | 0.427 | 0.047 | 0.002 | 0.038 | 0.02 | 0.546 | 0.102 | 0.145 | 0.018 | 0.518 | 0.065 | 0.109 | 0.532 | 0.083 | 0.127 | 0.467 | 0.428 | 0.045 |
| **Other Anti-hypertensive Use** | 0.032 | 0.028 | 0.02 | 0.292 | 0.03 | 0.053 | 0.059 | 0.052 | 0.322 | 0.061 | 0.084 | 0.008 | 0.264 | 0.002 | 0.025 | 0.272 | 0.01 | 0.033 | 0.262 | 0.239 | 0.023 |
| **Selective Serotonin Reuptake Inhibitor Use** | 0.187 | 0.113 | 0.46 | 0.052 | 0.001 | 0.027 | 0.075 | 0.286 | 0.239 | 0.188 | 0.16 | 0.356 | 0.164 | 0.113 | 0.086 | 0.508 | 0.46 | 0.435 | 0.051 | 0.079 | 0.028 |
| **Cardiovascular Medication Use** | 0.209 | 0.176 | 0.61 | 0.804 | 0.045 | 0.026 | 0.032 | 0.837 | 1.039 | 0.163 | 0.235 | 0.801 | 1.001 | 0.131 | 0.203 | 0.203 | 0.658 | 0.583 | 0.852 | 0.776 | 0.071 |
| **Other Cardiovascular Medication Use** | 0.144 | 0.047 | 1.495 | 0.683 | 0.027 | 0.025 | 0.097 | 1.308 | 0.54 | 0.171 | 0.17 | 1.434 | 0.637 | 0.074 | 0.073 | 0.66 | 1.528 | 1.527 | 0.708 | 0.707 | 0.001 |
| **Cardiac Medication Use** | 0.147 | 0.066 | 0.425 | 0.419 | 0.023 | 0.011 | 0.08 | 0.547 | 0.541 | 0.169 | 0.158 | 0.479 | 0.474 | 0.089 | 0.078 | 0.012 | 0.406 | 0.415 | 0.4 | 0.409 | 0.011 |
| **Implantable Cardioversion Defibrillator Placement** | 0.09 | 0.002 | 0.136 | 0.103 | 0.048 | 0.126 | 0.091 | 0.212 | 0.014 | 0.043 | 0.037 | 0.134 | 0.105 | 0.05 | 0.127 | 0.223 | 0.176 | 0.242 | 0.056 | 0.023 | 0.079 |
| **Pacemaker Placement** | 0.129 | 0.021 | 0.381 | 0.025 | 0.005 | 0.02 | 0.15 | 0.504 | 0.104 | 0.134 | 0.109 | 0.361 | 0.046 | 0.016 | 0.041 | 0.404 | 0.376 | 0.4 | 0.029 | 0.004 | 0.025 |
| **Coronary Arterial Bypass Graft Placement** | 0.129 | 0.037 | 0.038 | 0.327 | 0.032 | 0.117 | 0.165 | 0.091 | 0.451 | 0.097 | 0.244 | 0.075 | 0.291 | 0.069 | 0.081 | 0.364 | 0.006 | 0.155 | 0.359 | 0.211 | 0.149 |
| **Percutaneous Coronary Intervention** | 0.11 | 0.042 | 0.077 | 0.379 | 0.104 | 0.046 | 0.068 | 0.186 | 0.483 | 0.006 | 0.155 | 0.119 | 0.419 | 0.062 | 0.088 | 0.303 | 0.181 | 0.031 | 0.478 | 0.334 | 0.15 |
| **Prior Heart Failure Hospitalization** | 0.028 | 0.147 | | 0.805 | 0.01 | 0.15 | 0.175 | 1.514 | 0.837 | 0.019 | 0.122 | 1.253 | 0.642 | 0.157 | 0.297 | 0.506 | 1.485 | 1.712 | 0.816 | 0.977 | 0.14 |

| | | | | | | | | | | | | | | | | | | | | | |
|---|---|---|---|---|---|---|---|---|---|---|---|---|---|---|---|---|---|---|---|---|---|
| **Median Standardized Mean Difference (SMD)** | 0.144 | 0.075 | 0.219 | 0.214 | 0.048 | 0.058 | 0.074 | 0.212 | 0.171 | 0.079 | 0.118 | 0.224 | 0.178 | 0.05 | 0.041 | 0.203 | 0.2 | 0.164 | 0.184 | 0.166 | 0.028 |

**Table S5: Median Standardized Mean Difference of each covariate for pairs of TOPCAT participants and EHR patients used in this study analysis.** Abbreviations: ACE: Angiotensin Converting Enzyme, BH: Bridgeport Hospital, dL: deciliters, g:grams, GH: Greenwich Hospital, L: Liters, k: thousand, LMH: Lawrence & Memorial Hospital kg: kilograms, $m^3$: meters cubed, mg: milligrams, mmol: mili-moles, n: number, Q1: 25% percentile, Q3: 75% percentile, SMD: Standardized Mean Difference, SRC: St. Raphael's Campus, U: Units, uL: mililiters, YNHH: Yale New Haven Hospital, YSC: York Street Campus, Alk, Phos: Alkaline Phosphatase, ALT: Alanine Transferase, AST: Aspartate Transferase, BILITOT: Total Bilirubin, BMI: Body Mass Index, BUN: Blood Urea Nitrogen, CL: Chloride, CO2: Bicarbonate, DBP: Diastolic Blood Pressure, EF: Ejection Fraction, GFR: Glomerular Filtration Rate, GLU: Glucose, HCT: Hematocrit, HGB: Hemoglobin, HR: Heart Rate, K: Potassium, NA: Sodium, Plt: Platelets, SBP: Systolic Blood Pressure, WBC: White Blood Cells

| EHR patients with predicted spironolactone benefit on spironolactone with outcome (N, %) | EHR patients with predicted spironolactone benefit not on spironolactone with outcome (N, %) |
|---|---|
| 244 (20%) | 913 (14%) |

**Table S6: Incidence Rates of EHR Patients with Predicted benefit by TOPCAT-US participants from Spironolactone.** First column describes the number and percentage of outcome events in EHR patients with predicted spironolactone benefit on spironolactone, second column describes the number and percentage of outcome events in EHR patients with predicted spironolactone benefit not on spironolactone. Abbreviations: EHR: Electronic Health Record, N: Number

| EHR patients with predicted spironolactone benefit on spironolactone with outcome (N, %) | EHR patients with predicted spironolactone benefit not on spironolactone with outcome (N, %) |
|---|---|
| 34 (18%) | 122 (15%) |

**Table S7: Incidence Rates of EHR Patients with Predicted benefit by TOPCAT-Eastern Europe participants from Spironolactone.** First column describes the number and percentage of outcome events in EHR patients with predicted spironolactone benefit on spironolactone, second column describes the number and percentage of outcome events in EHR patients with predicted spironolactone benefit not on spironolactone. Abbreviations: EHR: Electronic Health Record, Ratio, N: Number

| | TOPCAT Participants missing values (N) | TOPCAT Participants missing values (%) | EHR Patients missing values (N) | EHR Patients missing values (%) |
|---|---|---|---|---|
| **ALBUMIN** | 179 | 5.20 | 2777 | 23.71 |
| **ALKPHOS** | 112 | 3.25 | 2769 | 23.64 |
| **ALT** | 42 | 1.22 | 2768 | 23.63 |
| **AST** | 47 | 1.36 | 2788 | 23.80 |
| **BILITOT** | 62 | 1.80 | 2766 | 23.62 |
| **BUN** | 773 | 22.44 | 2760 | 23.57 |
| **CL** | 124 | 3.60 | 2761 | 23.57 |
| **CO2** | 1603 | 46.53 | 2762 | 23.58 |
| **CREATININE** | 2 | 0.06 | 2758 | 23.55 |
| **GFR** | 2 | 0.06 | 7843 | 66.97 |
| **GLU** | 21 | 0.61 | 2793 | 23.85 |
| **HCT** | 27 | 0.78 | 761 | 6.50 |
| **HGB** | 23 | 0.67 | 805 | 6.87 |
| **K** | 4 | 0.12 | 2779 | 23.73 |
| **PLT** | 35 | 1.02 | 810 | 6.92 |
| **WBC** | 24 | 0.70 | 807 | 6.89 |
| **NA** | 6 | 0.17 | 5774 | 49.30 |
| **HR** | 5 | 0.15 | 32 | 0.27 |
| **SBP** | 4 | 0.12 | 29 | 0.25 |
| **DBP** | 4 | 0.12 | 36 | 0.31 |
| **BMI** | 10 | 0.29 | 297 | 2.54 |
| **Age** | 0 | 0.00 | 0 | 0.00 |
| **EF** | 1 | 0.03 | 0 | 0.00 |

**Table S8: Missingness of continuous variables in TOPCAT and the EHR patient datasets**. Abbreviations: ALKPHOS: Alkaline Phosphatase, ALT: Alanine Transferase, AST: Aspartate Transferase, BILITOT: Total Bilirubin, BMI: Body Mass Index, BUN: Blood Urea Nitrogen, CL: Chloride, CO2: Bicarbonate, DBP: Diastolic Blood Pressure, EF: Ejection Fraction, GFR: Glomerular Filtration Rate, GLU: Glucose, HCT: Hematocrit, HGB: Hemoglobin, HR: Heart Rate, K: Potassium, NA: Sodium, Plt: Platelets, SBP: Systolic Blood Pressure, WBC: White Blood Cells

| | Number of participants with missing values | Percent of participants with missing values (N=3445) |
|---|---|---|
| **Myocardial Infarction** | 3 | 0.09 |
| **Stroke** | 3 | 0.09 |
| **Chronic Obstructive Pulmonary Disease** | 3 | 0.09 |
| **Asthma** | 3 | 0.09 |
| **Hypertension** | 3 | 0.09 |
| **Peripheral Arterial Disease** | 3 | 0.09 |
| **Dyslipidemia** | 3 | 0.09 |
| **Atrial Fibrillation** | 3 | 0.09 |
| **Thyroid disease** | 3 | 0.09 |
| **Diabetes Mellitus** | 3 | 0.09 |
| **Smoker** | 4 | 0.12 |
| **Angiotensin Converting Enzyme- Inhibitor Use** | 3 | 0.09 |
| **Angiotensin Receptor Blocker Use** | 3 | 0.09 |
| **Beta Blocker Use** | 3 | 0.09 |
| **Calcium Channel Blocker Use** | 3 | 0.09 |
| **Diuretic Use** | 3 | 0.09 |
| **Glucose Lowering Medications Use** | 3 | 0.09 |
| **ACE-Inhibitor or Angiotensin Receptor Blocker Use** | 3 | 0.09 |
| **Aspirin Use** | 3 | 0.09 |
| **Nitrate Use** | 3 | 0.09 |
| **Statin Use** | 3 | 0.09 |
| **Warfarin Use** | 3 | 0.09 |
| **Lipid Lowering Medication Use** | 3 | 0.09 |

| | | |
|---|---|---|
| **Other Anti-hypertensive Use** | 3 | 0.09 |
| **Selective Serotonin Reuptake Inhibitor Use** | 3 | 0.09 |
| **Cardiovascular Medication Use** | 3 | 0.09 |
| **Other Cardiovascular Medication Use** | 3 | 0.09 |
| **Cardiac Medication Use** | 3 | 0.09 |
| **Anti-Hypertensive Medication Use** | 3 | 0.09 |
| **Non-Cardiovascular Medication Use** | 3 | 0.09 |
| **Implantable Cardioversion Defibrillator Placement** | 3 | 0.09 |
| **Pacemaker Placement** | 3 | 0.09 |
| **Coronary Arterial Bypass Graft Placement** | 3 | 0.09 |
| **Percutaneous Coronary Intervention** | 3 | 0.09 |

**Table S9: Missingness of categorical variables in TOPCAT trial**. Missingness of categorical variables was unable to be assessed in the EHR due to how the data is ascertained. Second column is the number of TOPCAT participants with the missing variable, and third column is the percentage of TOPCAT participant with the missing variable.
